# Using large language models to facilitate literature review and data extraction for infectious disease models: COVID-19 as a test case

**DOI:** 10.64898/2026.09.06.26362399

**Authors:** Xiyu Yang, Chui Yee Lee, Billy J Quilty, Linxi Zhang, Yao Mu, Mark Jit

## Abstract

Infectious disease transmission models are governed by parameters informed by systematic review of epidemiological literature. Large language models (LLMs) could facilitate this, but the reliability of the results to inform models has not been tested.

We built an open-source, end-to-end pipeline to simulate LLM performance in a hypothetical scenario where they were available to inform COVID-19 models developed during the first four months of 2020. It screened articles and extracted the reproduction number, serial interval, and incubation period from full-text PDFs. We applied it to 2,067 PubMed/medRxiv records published 31 December 2019–30 April 2020 using four models (GPT-5-mini, GPT-5.4, Claude Opus 4.8 and Gemini 2.5 Pro) and evaluated it against full-corpus human screening and 50-article extraction gold standards. We combined models post hoc, pooled the extracted values into an infectious disease (SEIR) model, and ran sensitivity analyses to test approaches to improve extraction accuracy.

Screening sensitivity was 0.72-0.95 and specificity 0.92-0.99. For articles reporting few values, extraction F1 was 0.90-0.96 with precision 0.91-1.00; across all articles, including those with dozens of stratified estimates, recall fell to 0.38-0.78. No fabricated values observed; errors were misassignments of values filed under the wrong parameter, or borrowed values treated as the study’s own. Incomplete extraction from dense articles was mainly due to prompting and output format, not model capability. Ensembling allowed recall-precision trade-offs, and correctness increased with model agreement, from about 30% at one vote to 92-95% at four. The pipeline processed the corpus in hours versus an estimated 130-265 person-hours of manual effort.

Our results show that current models can extract transmission parameters from unstructured literature accurately enough to inform outbreak modelling. The bottleneck lies in task specification, and careful prompt and output schema design are key to reducing misassignment errors. Human effort is best directed at workflow development and provenance validation.

## Introduction

Infectious disease models have informed public health responses during outbreaks, including COVID-19, mpox, and Ebola [1]. Early in an outbreak, these models provide valuable information to decision-makers about projected epidemic size, timing, and intervention impact. Their reliability depends on the epidemiological parameters that feed them [2,3]. Identifying these parameters relies on manual systematic review and data extraction of a literature base that is usually rapidly changing in a major outbreak or pandemic. Borah et al. estimated a mean of 67.3 weeks to complete and publish a systematic review [4]. It is also error-prone under time pressure, with Mathes et al. reporting data extraction error rates as high as 50% across four methodological studies [5].

Large language models (LLMs) may automate this evidence synthesis via an end-to-end pipeline for both literature screening and data extraction. LLMs have been used to assist screening of articles for inclusion in reviews [6,7], for full-text extraction of articles [8,9], and further downstream in transmission model code [10].

However, two issues remain unresolved before establishing LLMs as a routine work pipeline in a pandemic. First, LLM accuracy (agreement with human reviewers) varies across fields and tasks [6,11]. Concerns remain particularly around reliability, susceptibility to “hallucinations” (incorrect LLM responses generated as artefacts of the underlying model rather than a reflection of the data), and poor data provenance.

Second, few studies combine screening, extraction and model parameterisation in an automated, end-to-end pipeline, and many rely on older LLMs that may not reflect current capabilities. Existing methods were developed mainly for structured records and do not transfer cleanly to the epidemiological literature, which is heterogeneous in methods and population contexts. A pipeline for epidemiological parameter extraction has been introduced and evaluated against annotations covering 16,248 articles from systematic reviews of WHO priority pathogens, and it lost accuracy mainly at extraction [12]. That evaluation excludes SARS-CoV-2, and it stops at the extracted values without pooling them or carrying them into an epidemiological model. A similar pipeline on the WHO Collaboratory supports live outbreak response, but no formal accuracy evaluation had been published as of August 2026 [13].

The early COVID-19 literature offers an ideal test case because of the large volume of research that rapidly accumulated in early 2020 in preprints and journals [14]. We developed a semi-automated, open-source LLM pipeline that extracts three transmission parameters – reproduction number, serial interval, and incubation period – from the COVID-19 literature published in the first four months of 2020 to explore LLMs’ role in informing epidemiological models. The pipeline achieved high screening and extraction accuracy for most articles and produced epidemic curves comparable to those from human-extracted parameters, while the limitations observed in articles with dense parameters underscore the importance of workflow design and task specification.

## Methods

### Overview

We simulated a hypothetical modelling group on May 1, 2020, tasked with building a COVID-19 model from literature published through April 30, 2020, using today’s LLMs. We developed a semi-automated pipeline to search the COVID-19 literature, extract parameters relevant to epidemiological models and build a model (Fig 1). We built the pipeline iteratively on a development subset, then locked it and evaluated it against gold standards for screening, extraction, and downstream modelling. Screening criteria were taken directly from the protocol without tuning.

**Fig 1.**
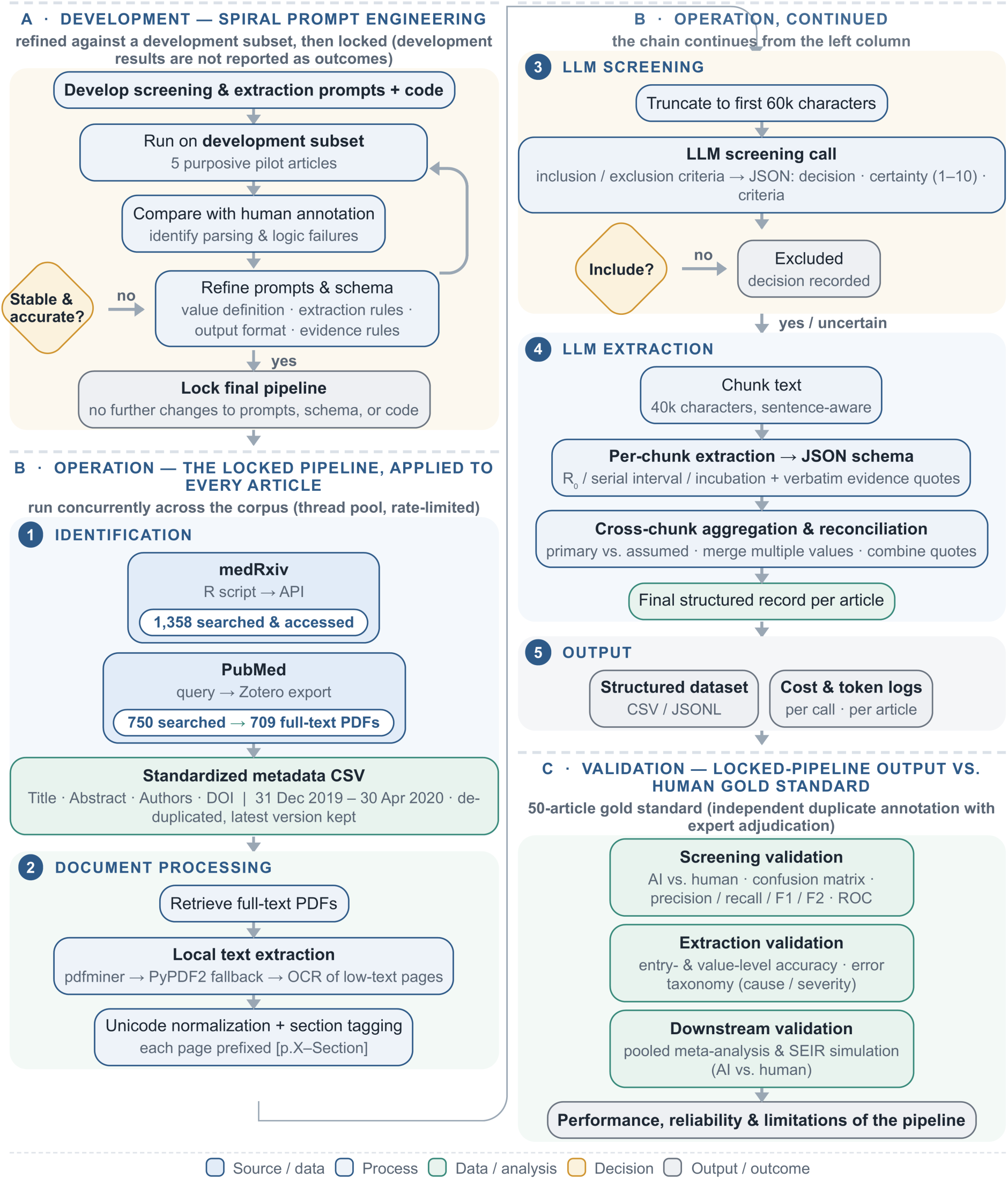
Development, operation and validation of the LLM-facilitated screening and extraction pipeline. Panels show (A) development and locking using five annotated articles, (B) application of the locked pipeline to the full corpus, and (C) evaluation against human screening and extraction reference standards, followed by pooled-estimate and SEIR analyses.

### Database search (Fig 1B, stage 1)

We systematically searched medRxiv and PubMed for articles from December 31, 2019, through April 30, 2020. We searched both databases because medRxiv preprints hosted the majority of early parameter estimates whereas peer review later reshaped the report content [15]. Duplicates were removed; for preprints, only the latest version within the date cutoff was retained. The search strategy combined pathogen and parameter terms (S1 Appendix, Section 1). medRxiv articles were retrieved via an R script utilizing its API; PubMed records were exported from Zotero, with articles requiring subscription accessed manually. S1 Appendix, Section 4 gives the modified PRISMA for flow of records through the process.

### Human gold standard (Fig 1C)

We established a gold standard (i.e. high-reliability dataset for verification) by human screening and full-text extraction. A single reviewer categorized articles as “include,” “uncertain,” or “exclude”, with uncertain articles re-adjudicated by a blinded expert reviewer. We excluded animal-only studies, case reports, non-original reports, articles without numerical parameter estimates, and purely theoretical modelling papers that neither estimated a target parameter nor used a published one as an input. Human screening used titles and abstracts first and consulted the full text where that left the decision unclear, while the models screened on the article text directly. Full inclusion and exclusion criteria are in S1 Appendix, Section 2.

For full-text extraction, we randomly sampled 50 articles that passed human screening (25 from PubMed, 25 from medRxiv) and manually extracted the three transmission parameters (if available). Two researchers with epidemiological training (blinded to LLM output) independently extracted parameters from each article, following the protocol in S1 Appendix, Section 3. Discrepancies between reviewers were reconciled through discussion and, where consensus could not be reached, adjudicated by a senior epidemiological modeller to yield final consensus.

Given the predominance of not-reported entries, we quantified inter-rater reliability between the two reviewers using Gwet’s AC1 for reportability (whether a parameter was recorded for an article) [16] and Lin’s concordance correlation coefficient (CCC) for numeric values [17].

### LLM-facilitated pipeline (Fig 1A and 1B, stages 2 to 5)

We iteratively refined prompts on a 5-article pilot set (none in the gold standard) [18,19]. The LLMs output extracted data in JSON format [20] requiring at least one verbatim quotation per value and a maximum of three supporting quotations per parameter. Prompts are in S2 Appendix, Section 2. We used four LLMs (GPT-5.4, GPT-5-mini, Claude Opus 4.8, Gemini 2.5 Pro) in a Python pipeline that processed each article through document processing, screening, extraction (with cross-chunk aggregation), and structured output (Fig 1B; details in S2 Appendix, Section 1).

### Sensitivity analysis

We conducted sensitivity analyses varying output format, instruction wording, input mode (text vs. page images), and merging rule on five dense article-parameter pairs. A set of small-granularity articles was then evaluated to test for potential adverse effects of these varied factors on typical articles. Full sensitivity analysis descriptions and results are in S4 Appendix.

### Evaluation metrics and statistical analysis

We compared model outputs to the gold standard by computing true positives (TP), false positives (FP), true negatives (TN), and false negatives (FN) for both stages. For screening, counts were at the article level.

An entry is one article-parameter pair; a value is one number within an entry. Extraction accuracy was evaluated at the parameter level end-to-end, so an error was recorded for all parameters in a paper that a model failed to shortlist at screening. A TN was recorded when a model correctly left a non-reported parameter as “N/A.”

From these counts, we computed recall, precision, specificity, accuracy, and F-measures (definitions in S5 Appendix). F2, which weights recall above precision, was used because a missed estimate is more costly than a spurious one. For the vote-count ranking in the ensemble analysis we also report the area under the receiver operating characteristic curve (AUC), which summarizes discrimination across all decision thresholds, from 0.5 (chance) to 1.0 (perfect).

Uncertainty was quantified with 95% confidence intervals. Because individual extracted values are nested within articles, we used a cluster bootstrap resampling articles rather than values to avoid falsely narrowing intervals for articles reporting many values. S5 Appendix states the interval method for each metric. We compared models using McNemar’s test (screening) and paired cluster-bootstrap (extraction).

### Error classification

We classified each LLM error (discrepancy between model output and the human gold standard) as false positive or false negative and assigned it to one of three categories: (1) identity/provenance confusion, (2) granularity omission, or (3) screening exclusion (S3 Appendix, Section 1).

### Pooled estimates and heterogeneity

We pooled LLM- and human-extracted parameters separately using inverse-variance weighted random-effects models. When a study reported multiple estimates, each value was weighted by 1/n (n = the number of values from that study). Only studies that reported 95% CIs/CrIs/eCIs or SDs (converted to 95% CI as mean ± 1.96 × SD) were included in the pooled estimates. Two systematic reviews/meta-analyses were excluded to avoid double-counting primary research [21,22]. Statistical heterogeneity between studies was assessed using between-study heterogeneity variance, τ^2^, the I^2^ statistic and Cochran’s Q statistic. Meta-analysis and heterogeneity estimation were performed in R 4.6.0 using the meta package.

### Transmission model simulation

We constructed an SEIR (Susceptible-Exposed-Infected-Recovered) transmission dynamic model with homogeneous mixing and a closed population to simulate SARS-CoV-2 transmission (S6 Appendix, Section 1). This was parameterised separately with the parameters extracted by LLMs and humans. To capture uncertainty, we bootstrapped R0 and incubation period 50 times each, independently from each LLM- and the human-extracted distribution. The bootstrapping used the full distribution of extracted parameter values except one extreme R0 value at 59.3 (that was correctly identified by both LLMs and humans). Only basic reproduction numbers were included; Rt and Rc were excluded. Serial interval was evaluated at extraction but not used in the simulation. The sampled incubation period was used to calculate the rate of progression to infectiousness σ, which is 1/incubation period. The infectious period was set to 5 days [23], and the recovery rate γ, was computed as 1/5. Birth and mortality were not included. We summarized the simulated epidemic curves as mean ± standard deviation (SD), and functional median and pointwise median with inter-quartile range (IQR). The SEIR model was developed in R 4.6.0.

### Multi-model ensemble

We combined outputs from the four models using multi-model agreement as a tunable decision rule. For screening, we combined each article’s four include/exclude votes (“uncertain” counted as include) using a k-of-N threshold: retain if at least k of four models vote to include. For extraction, we matched the values reported in each model’s summary field (a point estimate with its reported uncertainty) across models by exact numbers, and each distinct value received a vote count (number of models reporting it). The ensemble at threshold k retains the values with support ≥ k. Both ensembling steps were post-processing of the locked outputs and added no model calls.

### Ethics statement

This study analysed published scientific articles and preprints. It involved no human participants, no animals, and no personally identifying data, so ethical approval was not required.

### Use of AI tools

Four large language models (GPT-5.4, GPT-5-mini, Claude Opus 4.8, and Gemini 2.5 Pro) are the subject of this study, with their use and evaluation described in the article. Separately, Claude was used as an assistive tool to debug Python and R code, to suggest writing improvements in the manuscript, and to check for formatting consistency. All AI-assisted outputs were reviewed, verified, and edited by the authors before inclusion. All hypotheses, interpretations, results, conclusions and limitations reported here reflect the original work of the authors.

## Results

### Literature search and automated screening performance

All four models performed well on identifying articles from medRxiv, with trade-offs between precision and recall (Fig 2 and S7 Appendix, Table A). Performance on PubMed was lower overall than on medRxiv. GPT-5-mini recovered more relevant articles (higher recall, fewer false negatives) but had lower precision and specificity; GPT-5.4 was more conservative and precise. On PubMed, Gemini 2.5 Pro achieved the highest F1 (0.885) while GPT-5-mini achieved the highest sensitivity (0.939) and Claude Opus 4.8 the lowest (0.798). On medRxiv articles, Claude Opus 4.8 performed best for precision and specificity. Under F2 (which weights recall above precision) GPT-5-mini performed best on both datasets, outperforming its larger counterpart (GPT-5.4).

**Fig 2.**
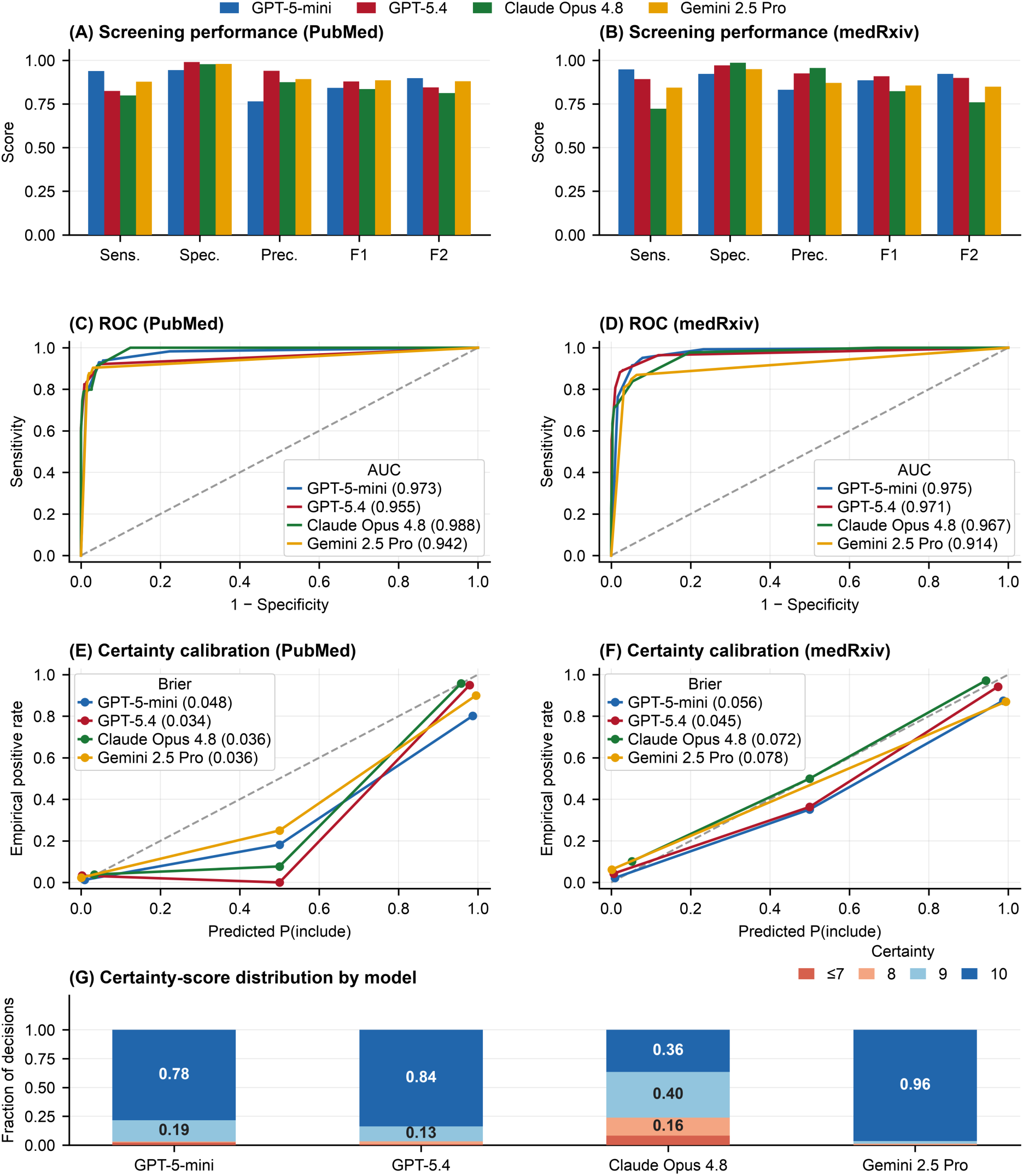
Screening performance, ROC curves, certainty calibration and certainty-score distributions across models and datasets. Panels (A, C, E) present PubMed results and panels (B, D, F) present medRxiv results; panel (G) pools certainty scores across both datasets by model. (A, B) show sensitivity, specificity, precision, F1 and F2 against human reference labels. (C, D) show receiver operating characteristic curves; dashed diagonals indicate chance discrimination and area-under-the-curve values are reported in the legends. (E, F) show observed inclusion rates against inclusion probabilities derived from each model’s decision and certainty score; dashed diagonals indicate perfect calibration and the legends report Brier scores. AUC, area under the curve; ROC, receiver operating characteristic.

The receiver operating characteristic (ROC) curves confirm strong, comparable discrimination for all models on both databases: every curve rises steeply to high sensitivity at a low false-positive rate, and all eight areas under the curve exceed 0.91, with Claude Opus 4.8 attaining the highest AUC on PubMed and GPT-5-mini the highest on medRxiv.

We next examined each model’s self-reported certainty score (1-10) against its actual decision accuracy (Fig 2, panels E and F). Calibration was reasonable overall (per-corpus Brier 0.034-0.078; expected calibration error 0.011-0.078), but the four models used the scale very differently (Fig 2, panel G): GPT-5-mini and GPT-5.4 gave most decisions at the maximum certainty of 10, Gemini 2.5 Pro was almost saturated (mean certainty 9.95, nearly all decisions at 9-10) and was the least calibrated, whereas Claude Opus 4.8 spread its decision certainty scores from 2 to 10 and was the only meaningfully discriminating score. The models differed in how confidently they made false-negative screening errors. For GPT-5-mini, GPT-5.4, and Gemini, wrongly excluded articles were typically issued with high certainty (median 9-10), so the self-reported score could not flag potential misses. Only Claude concentrated errors at low certainty, and routing its decisions at certainty ≤7 to a human reviewer would recover 44% of its missed articles (58 of 131) at an 8% review burden (Fig 3A).

**Fig 3.**
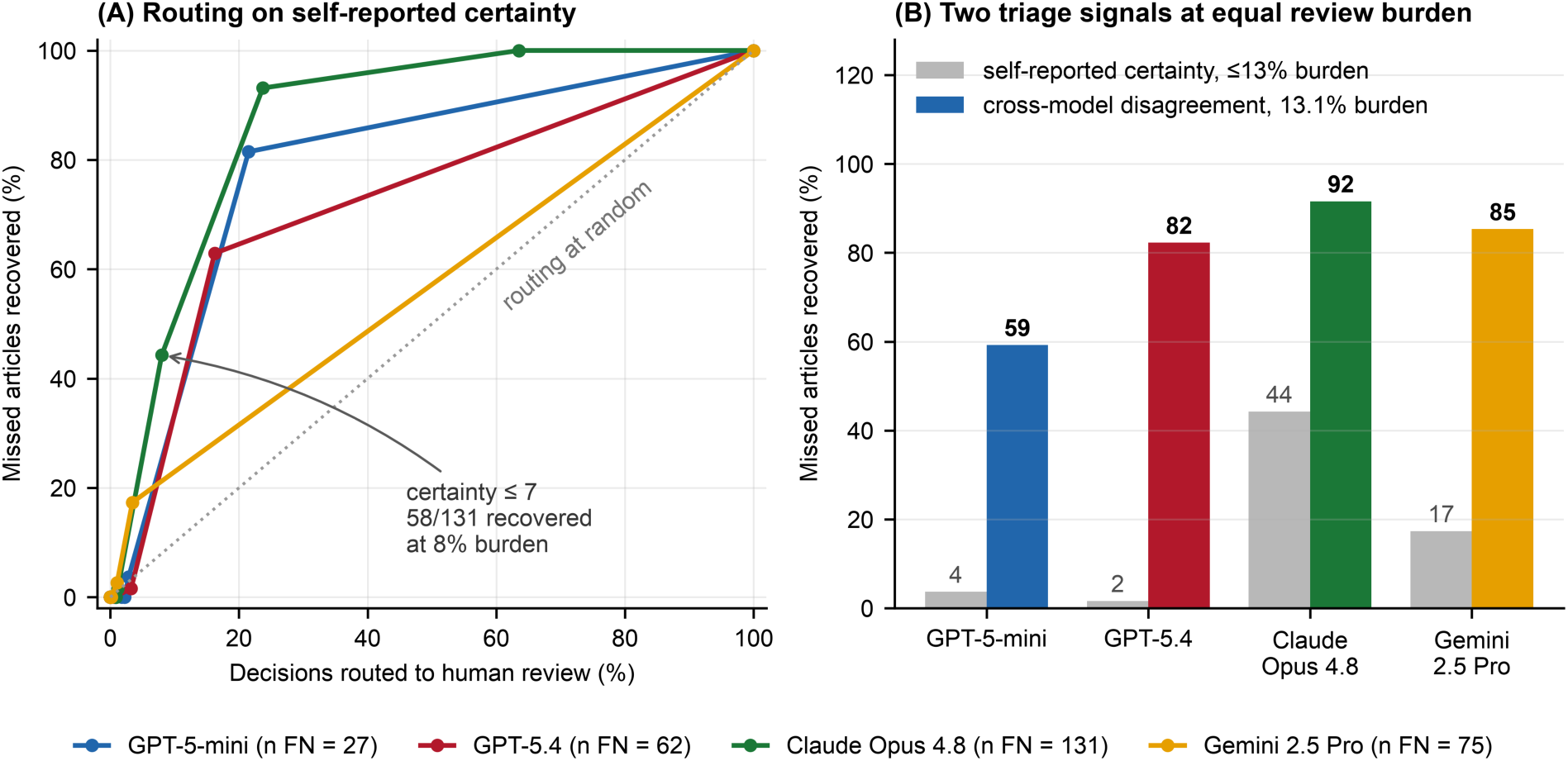
Recovering missed articles by routing decisions to a human reviewer. (A) Percentage of a model’s wrongly excluded articles recovered against the percentage of all decisions routed, sweeping the self-reported certainty threshold from 1 to 10; the dotted diagonal is routing at random. (B) The same recovery at a matched review burden, comparing routing on self-reported certainty against routing on cross-model disagreement, which requires no model to calibrate itself. Both panels assume a reviewer resolves every routed article correctly and are therefore upper bounds.

### Full-text data extraction accuracy

The two human data extractors agreed on reportability in 93% of entries (AC1 0.85), and almost perfectly on the value in the 36 entries where both recorded a number (CCC 0.961; within ±5% in 92%). Only ∼9% of entries required adjudication by the senior modeller. All four LLMs showed high, tightly clustered extraction accuracy.

On articles each model carried forward (excluding those with dozens of stratified values), extraction performance was tightly clustered: F1 0.90–0.96, recall 0.84–0.96, precision 0.91–1.00. Claude Opus 4.8 returned no false positives on PubMed. Including screening losses, recall dropped to 0.65–0.92 with precision unchanged because screening misses add false negatives but not false positives. The largest drop was Claude Opus 4.8 on PubMed (0.919 to 0.654). Across the full gold standard (including dense articles), recall fell to 0.38–0.78 (Fig 4A, B). Per-model values for all three scopes are in S7 Appendix, Table E.

**Fig 4.**
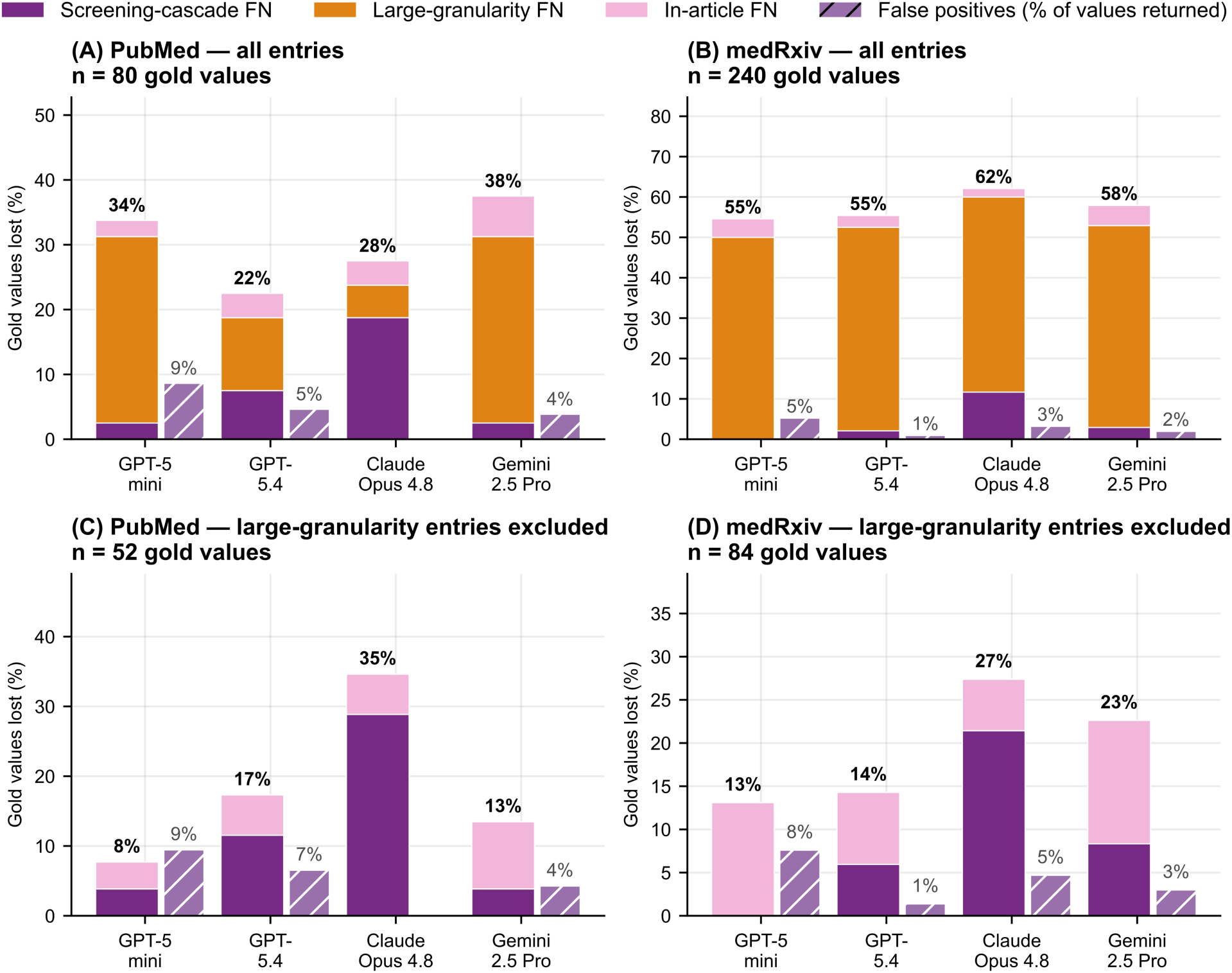
Extraction-error decomposition by model and corpus. Panels (A, B) show all error entries; panels (C, D) show the same with large-granularity entries excluded, for PubMed and medRxiv respectively. Stacked bars give false negatives as a percentage of the gold-standard values in that panel (80 and 240 in A and B; 52 and 84 in C and D). The segments are screening-cascade false negatives, large-granularity false negatives, and in-article false negatives from identity and provenance confusion. The hatched bar gives false positives as a percentage of the values a model returned, plotted separately because the denominator differs.

### Error classification and root cause analysis

We observed no fabricated values across any model or database. Every erroneous output corresponded to a real number in the source text, misassigned to a different parameter or source. Error patterns were consistent across models (S3 Appendix, Section 2). Fig 4 shows per-model frequencies; S3 Appendix, Table A lists representative cases.

Manual review traced these errors to the tendency of models to maximise recall during extraction by surfacing any value that appeared relevant even if it is peripheral, secondary, or from a different pathogen. The same tendency produces the false-positive screening errors (S3 Appendix, Table C), where the mere presence of a transmission-parameter number, even one that was cited, definitional, or purely descriptive, was enough to include an article. Because models record these values’ status in the summary, misassigned values can be recovered by a downstream filter without re-reading the source (S3 Appendix, Table B; S8 Appendix).

### Pooled parameter estimates and transmission model

Across all three parameters, every LLM pooled estimate was within 15% of the gold standard (Table 1; S6 Appendix, Section 2 for scatter plots; S6 Appendix, Fig C for raw distributions). Heterogeneity was consistently high for R_0_ and incubation period across both the LLM- and human-extracted pools (all I^2^ > 90%). 59% and 56% of R_0_ and incubation period estimates came from just two papers, so this heterogeneity partly reflects within-study variation across scenarios in the same study. For the serial interval, all estimates came from just two to three studies.

**Table 1.** Pooled estimates and statistical heterogeneity for extracted parameters. Pooled estimates were calculated using an inverse-variance weighted random-effects model. Heterogeneity between studies was assessed using the τ^2^ (tau squared, the between-study variance), the I^2^ heterogeneity statistic, and Cochran’s Q test. CI: confidence interval; df: degree of freedom.

| | Pooled estimates | Discrepancy (gold vs. LLMs) | 95% CI | $\tau^2$ | I<sup>2</sup> score (%) | *p*-value (Q of heterogeneity; df) | Number of studies |
| --- | --- | --- | --- | --- | --- | --- | --- |
| Basic reproduction number, R<sub>0</sub> |  |  |  |  |  |  |  |
| Gold standard | 3.36 | - | 3.17 to 3.54 | 0.76 | 96.9 (96.6-97.1) | 0.000 (4722.24; 148) | 11 |
| GPT-5.4 | 2.94 | 0.42 | 2.55 to 3.32 | 0.98 | 99.0 (98.9-99.1) | 0.000 (4803.91; 49) | 11 |
| GPT-5-mini | 2.91 | 0.45 | 2.42 to 3.40 | 1.30 | 99.3 (99.2-99.3) | 0.000 (4437.48; 33) | 11 |
| Claude Opus 4.8 | 2.97 | 0.39 | 2.48 to 3.46 | 1.31 | 99.2 (99.1-99.3) | 0.000 (4432.55; 35) | 11 |
| Gemini 2.5 Pro | 2.88 | 0.48 | 2.42 to 3.35 | 1.20 | 99.3 (99.2-99.4) | 0.000 (4847.41; 33) | 11 |
| Incubation period |  |  |  |  |  |  |  |
| Gold standard | 6.99 | - | 6.18 to 7.78 | 2.24 | 91.2 (88.3-93.4) | 0 (273.6; 24) | 6 |
| GPT-5.4 | 6.93 | 0.06 | 5.40 to 8.46 | 6.77 | 94.7 (92.9-96.0) | 0 (319.2; 17) | 8 |
| GPT-5-mini | 6.92 | 0.07 | 5.39 to 8.45 | 7.54 | 96.4 (95.3-97.3) | 0 (419; 15) | 6 |
| Claude Opus 4.8 | 6.30 | 0.69 | 5.10 to 7.51 | 4.37 | 95.5 (93.9-96.7) | 0 (312.74; 14) | 8 |
| Gemini 2.5 Pro | 6.59 | 0.4 | 5.22 to 7.97 | 5.57 | 95.9 (94.4-97.1) | 0 (296.15; 12) | 6 |
| Serial interval |  |  |  |  |  |  |  |
| Gold standard | 6.53 | - | 5.62 to 7.45 | 1.86 | 53.7 (18.4-73.7) | 0.0057 (32.39; 15) | 2 |
| GPT-5.4 | 5.83 | 0.7 | 5.30 to 6.36 | 0 | 0 (0-62.4) | 0.839 (4.94; 9) | 3 |
| GPT-5-mini | 5.83 | 0.7 | 5.30 to 6.36 | 0 | 0 (0-64.8) | 0.757 (5.01; 8) | 3 |
| Claude Opus 4.8 | 6.63 | -0.1 | 5.13 to 8.13 | 3.31 | 79.5 (60-89.5) | 0.000 (34.09; 7) | 3 |
| Gemini 2.5 Pro | 5.69 | 0.84 | 5.14 to 6.24 | 0 | 22.8 (0-66.9) | 0.263 (6.47; 5) | 2 |

Fig 5 shows curves simulated from an SEIR model parameterized with bootstrapped samples of the extracted R_0_ and incubation period values. The functional median represents the single bootstrapped curve with the lowest global distance, whereas the pointwise median is computed independently at each time step across the 50 simulations. LLM- and human-extracted parameters produced broadly overlapping epidemic curves. Peak timing and peak infected proportion overlapped across all sources including the gold standard, alongside visible differences in peak magnitude.

**Fig 5.**
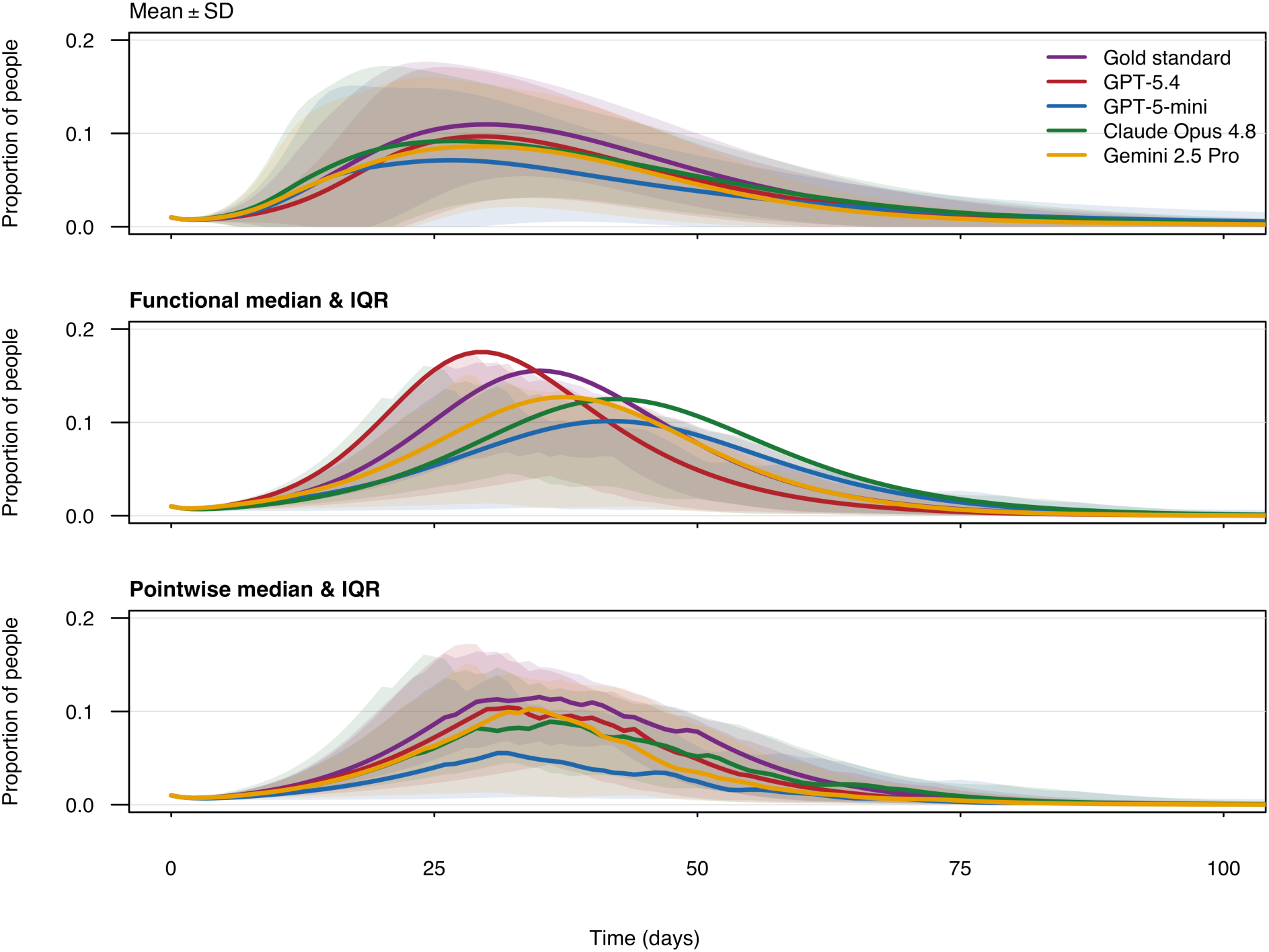
Simulated COVID-19 epidemic curves from human- and LLM-extracted parameters. Colours denote the source of the parameters: human (purple), GPT-5.4 (red), GPT-5-mini (blue), Claude Opus 4.8 (green) and Gemini 2.5 Pro (yellow). Lines are the pointwise mean (top), functional median (middle) and pointwise median (bottom) of the proportion infected over time. Shaded regions show the SD (top) and interquartile range (middle, bottom). R_0_ and the incubation period were independently sampled with replacement from the extracted values (n=50 bootstrap iterations), with the infectious period fixed at 5 days.

### Efficiency and cost analysis

Processing the corpus, screening every record and extracting full text from those that passed, for the 2,065 records carrying complete cost logs, required 29–40 million tokens per model (approximately 14,000–19,000 tokens per article) (S9 Appendix, Table A; Fig A, panel B). Median per-article latency was 3-11 seconds (S9 Appendix, Fig A, panel A). Approximate API costs, anchored to billed spend and reported only for scale, ranged from about $25 for the deployed GPT-5-mini to about $175–240 for Claude Opus 4.8; the deployed model cost roughly $0.014 per medRxiv article and $0.008 per PubMed article.

For human extraction, on the 50 timed gold-standard articles, the two annotators averaged 9–25 minutes per article (medRxiv 14.4 and 24.5 minutes; PubMed 8.9 and 19.6 minutes). Because a human extracts only the records that pass screening rather than the full retrieval, manual extraction of the 503 screened-in records would require an estimated 110–196 person-hours across the two annotators; adding manual screening of all records (at an assumed 0.5–2 minutes per abstract) gives an estimated end-to-end human effort of roughly 130–265 person-hours. The developed pipeline completed the same task in a few hours.

### Multi-model ensembling

The four-model vote threshold k allows recall and precision to be traded off against each other (S7 Appendix, Table C). The union (k ≥ 1) maximized sensitivity (0.965 on PubMed, 0.982 on medRxiv) and the recall-weighted F2 score (0.900 and 0.940), exceeding every individual model, whereas the unanimous rule (k ≥ 4) maximized precision and specificity (precision 0.976 and 0.993). The intermediate k ≥ 2 rule gave the best balanced F1 (0.904 and 0.893), matching the strongest single model’s F1 at higher precision. The ensemble’s vote-count ROC AUC (0.993 on PubMed, 0.987 on medRxiv) exceeded that of all four individual models on both corpora. Disagreement among the four models localises the screening misses. The 270 articles on which the vote was not unanimous, 13.1% of the corpus, contain 92% of the articles Claude Opus 4.8 wrongly excluded, 85% of Gemini 2.5 Pro’s, 82% of GPT-5.4’s and 59% of GPT-5-mini’s. Routing on each model’s own certainty score at the same review burden reaches 44%, 17%, 2% and 4% respectively (Fig 3B). Both are upper bounds, since each assumes a reviewer resolves every routed article correctly.

Value-level support voting behaved analogously at extraction (S7 Appendix, Table D, Fig 6A): the union of the four models’ values recovered more gold values than the best single model (recall 0.981 against 0.885 on PubMed; 0.845 against 0.750 on medRxiv), and raising the threshold traded recall back for precision (0.92 to 0.95 at k ≥ 4). The per-value support count behaved in this dataset as an agreement-based confidence signal that no single model can supply (Fig 6B). Values reported by all four models matched the gold standard in 92% (PubMed) and 95% (medRxiv) of cases, falling monotonically to 29% and 32% for values reported by only one.

**Fig 6.**
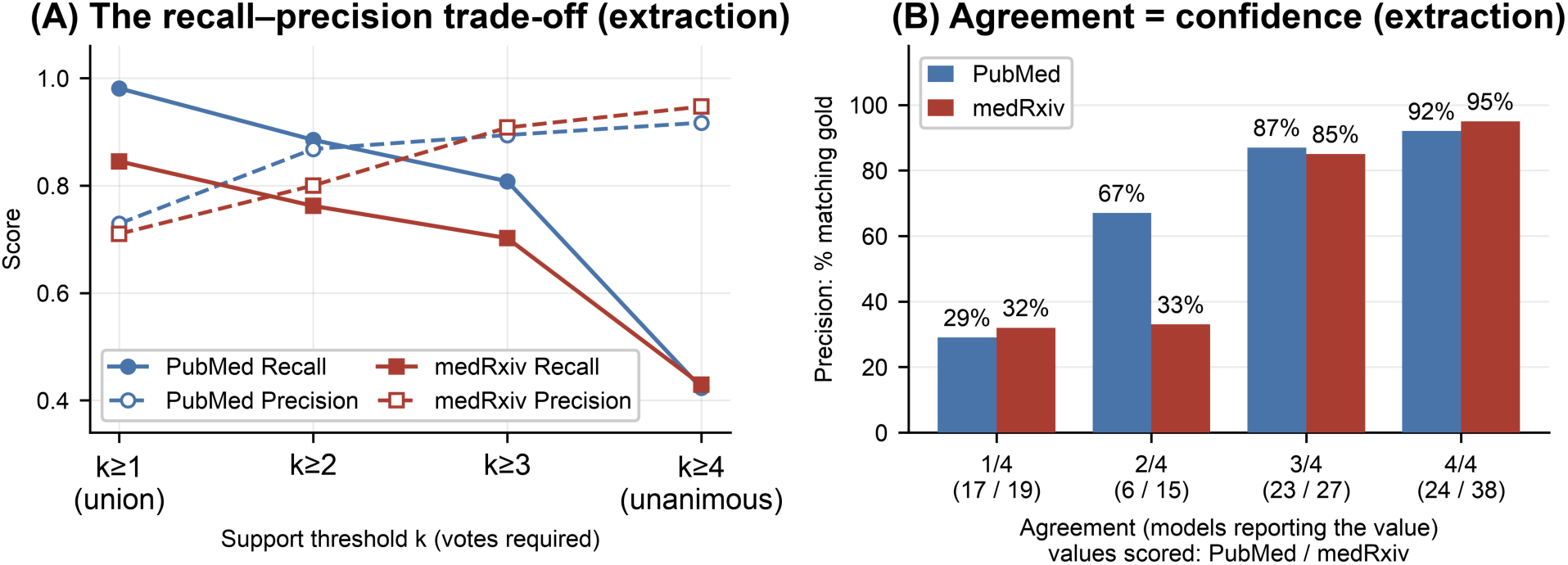
Multi-model ensemble at extraction. (A) The recall–precision trade-off: as the support threshold k rises from the union (k ≥ 1) to unanimous (k ≥ 4), recall falls and precision rises. (B) Agreement as confidence: the precision of an ensemble value (its probability of matching the gold standard) increases monotonically with the number of models that separately report it, from ≈ 30% at 1/4 vote to 92–95% at 4/4 votes. Large-granularity entries are excluded throughout.

The union’s recall advantage came almost entirely from model-specific errors that other models avoided; it did not recover the omissions (see S4 Appendix, Section 2) which all four models miss. When large-granularity entries were retained the union’s recall fell to 0.46 on medRxiv, indicating that ensembling raises the performance floor set by independent errors but not the ceiling set by the errors the models share.

## Discussion

### Principal findings

This study evaluated whether LLMs can accelerate parameter identification and extraction for real-time epidemiological models during outbreaks. LLMs were able to reproduce human screening and parameter extraction with high accuracy for articles reporting one or a few values per parameter (Fig 4C, D; S7 Appendix, Table E), but accuracy fell for articles with many stratified values. Most errors were attributable to prompt ambiguity and could have been avoided with further refinement. We deliberately limited prompt and pipeline refinement after initial testing to simulate real-world time pressure, under which teams working in situations such as a pandemic may have limited opportunity to iteratively validate and improve their approach. This also reflects the practical constraints of systematic review methodology, where substantive changes to the review protocol may require repeating affected stages of the search and analysis.

Hallucination did not appear despite being a concern around LLMs in the literature. All four LLMs performed reasonably well, but an ensemble of the models gave the best overall performance.

### Relation to prior work and what this study adds

Previous proof-of-concept work has established that LLMs can approach human accuracy on structured data extraction for evidence synthesis [8], with later studies improving performance through prompt engineering [19], domain-specific schemas [24], and collaborative multi-reviewer emulation [9]. Multi-model comparisons found that accuracy varies by model and task, and errors accumulate once single values are aggregated [25,26]. A benchmark of three models across three medical domains found high precision with recall lost to omission, with recall increased by up to 15% with prompts customised to the task [27]. ExtractBench reports a similar pattern outside biomedicine: where models returned valid output, they were correct on about 73% of fields, but on a 369-field schema no model returned valid output at all. The authors attribute this to the output volume required from the models rather than to input length [28].

In epidemiology, Padarha et al. evaluated a pipeline like ours against expert annotations from systematic reviews of WHO priority pathogens [12]. They found, as we did, that extraction rather than screening is where accuracy is lost, with no model exceeding an average field-level F1 of 0.67. Their prompt adopted the output format asking for one row per reported estimate, showing that this scheme alone does not guarantee high accuracy. Our comparison suggests the effect of the output format, as changing it alone without changing the model or the articles recovered most of what the deployed pipeline had missed. Their reference standard covers Ebola, Lassa, SARS-CoV-1 and Zika, but not SARS-CoV-2, and they do not evaluate pooling across studies. Sujau et al. ranked candidate infectious disease studies without extracting parameter values [29].

### Practical guidance for specifying a workflow

We found that LLM performance was critically dependent on the prompt used and the wider pipeline developed for the task. A workflow based on the lessons we learnt from this activity is summarised below. S8 Appendix lists implemented elements and recommendations from the error analysis.

##### Box 1. Practical guidance for specifying an LLM-facilitated systematic review workflow to parameterise an infectious disease model

**1. Define the prompt and schema before building the pipeline.** Invest the most effort in prompt and schema development early in the process. Many of the failure modes we discovered after deployment could have been discovered during initial development. The task specification should be designed specifically for the requirements of extracting parameters for infectious disease models, rather than using a generic extraction prompt.
**2. Optimize screening for recall.** Models should be prompted to include an uncertain category when classifying articles being screened, and then to route these cases for human review. If multiple models or agents are deployed, then disagreement between them can also be used as a trigger for human review. State provenance rules explicitly, including how to distinguish a study’s own estimates from borrowed or assumed values.
**3. Design the schema around individual values.** Give each extracted value its own row, and provide fields for all relevant parameters rather than forcing unfamiliar parameters into related fields. For example, generation time should not be recorded as a serial interval simply because no separate field exists. Include structured fields for provenance, such as study-derived, borrowed, or assumed, rather than relying only on free text.
**4. Keep prompt rules clear and consistent.** Make instructions specific, actionable, and consistent across pipeline stages. Avoid arbitrary limits to extraction values that can cause omissions in articles reporting many values. Build the development set to include typical, dense, ambiguous, and edge-case articles so that prompt and schema failures are found before full-scale deployment.
**5. Reassess pipeline mechanics as LLMs improve.** Several constraints we faced such as separate character recognition of optically scanned articles, character limits in context windows, article segmentation and cross-segment reconciliation are much less relevant for more recent LLMs. The rationale for pipeline design choices needs to be documented so they can be reassessed as model capabilities change.
**6. Use model ensembles to control recall and precision.** Use a k-of-N agreement threshold as a tunable recall–precision control for screening and extraction. The number of models reporting the same value can be used as a confidence signal for human review. However, ensembling does not fix systematic omissions or specification problems, so well-designed schema and prompts are still needed.
**7. Analyze disagreements, not just accuracy.** Examine disagreements between models (or between a model and a human review) to determine whether they reflect genuine errors or reasonable boundary disagreements.

## Strengths, limitations and recommendations for future work

Our study has several strengths. To our knowledge, it is the first to evaluate LLM-extracted transmission parameters through pooled meta-analysis informing an infectious disease model, showing that LLM-derived parameters produce epidemic curves comparable to human-extracted ones. The broadly favorable results suggest that LLMs could enhance modeling efforts to inform responses to a range of future outbreaks, saving time and human resources during these critical events. We selected four models spanning a wide capability and cost range, allowing us to separate model-specific differences from shared failure modes and test ensembling as a quality-control tool. Every screening decision was traceable to the rule that produced it, so misses are not attributed to general model capability. Finally, our post-hoc sensitivity analysis attributed recall loss in dense articles mainly to output schema, pointing to a practical but underused diagnostic for LLM pipeline failures.

Our study also has several limitations. The extraction evaluation relied on 50 double-annotated articles and 150 entry-level comparisons; a larger sample would tighten the accuracy estimates. The LLMs were likely pre-trained on past COVID literature and its meta-analyses, so we cannot exclude the possibility that they produced output based on their training data rather than the papers provided to them, although we minimized this by directing them to extract values from papers provided.

Finally, the four models represent a snapshot of a fast-moving field, and newer versions have since been released. However, the failure modes were shared across models ranging from a small, inexpensive model to three frontier systems. Hence, we expect our key findings to remain valuable even as LLMs advance.

A natural extension is to move from a fixed, single-pass pipeline toward agentic architectures, in which autonomous language model agents plan, use external tools, and critique their own output across several steps. The field is already moving this way [30], from multi-model workflow that emulates independent reviewers [9] and cross-checking between models [31] to retrieval-augmented pipelines [7] and guardrailed extraction with validation and retry [32]. Such a system could enforce the provenance and pathogen rules that a single-pass prompt leaves to the model’s discretion, re-read the articles that a single screening criterion currently excludes, and digitize figures where a value is legible only in a plot (S8 Appendix).

Authors of outbreak reports can improve machine extractability with clearly labeled tables, explicit uncertainty measures, and primary estimates separated from assumed or cited values. If LLMs become routine for extracting data from the literature, articles in computationally intensive fields like infectious disease modelling may increasingly be written for both human and machine readability.

## Acknowledgments

We thank Jun Shern Chan for insightful comments and suggestions that helped us situate the study within current developments in artificial intelligence.

## Data and Code Availability

Code is publicly available at https://github.com/MarkYXY05/covid-lit-llm-code. The data underlying the reported findings are publicly available at https://github.com/MarkYXY05/covid-lit-llm-data. Final data package will be archived in Zenodo under the reserved DOI https://doi.org/10.5281/zenodo.22267835 before journal publication.

## Supporting information

**S1 Appendix. Search strategy and screening protocol.** Full PubMed and medRxiv search strings, the inclusion and exclusion criteria applied at screening, and the search-retrieval and gold-standard procedure.

**S2 Appendix. Pipeline implementation and prompts.** Model configuration and the system and user prompts exactly as deployed.

**S3 Appendix. Error taxonomy, observed patterns and representative cases.** Definitions of the three error categories, how each appeared in the corpus, the screening losses attributable to the screening approach, and worked examples of extraction errors, constructive provenance labelling, and screening errors.

**S4 Appendix. Sensitivity analysis: output format, input mode and aggregation.** Design and results of the dense-target factorial experiment, and the evidence that granularity omission is a property of the output format.

**S5 Appendix. Evaluation metrics and interval construction.** Definitions of every reported metric and the interval method used for each.

**S6 Appendix. Transmission model and downstream comparison.** SEIR model structure and equations, and the comparison of AI- and human-extracted values.

**S7 Appendix. Full performance tables.** Confusion counts for screening and extraction, screening performance with confidence intervals, the two ensemble tables, and value-level extraction quality by scope.

**S8 Appendix. Operational guidance and what we tested.** Each recommendation labelled by whether it formed part of the locked pipeline or arises from the error analysis.

**S9 Appendix. Efficiency and cost.** Token counts, per-article time and approximate API cost by model, with the two human annotators for comparison.

S10 Checklist. PRISMA 2020 checklist.

## References

1. Rao D, Tanveer A, Iftekhar EN, Müller SA, Sherratt K, Röbl K, et al. The utility of infectious disease modelling in informing decisions for outbreak response: A scoping review. PLOS Global Public Health. 2025;5: e0005120. doi:10.1371/journal.pgph.0005120

2. Heesterbeek H, Anderson RM, Andreasen V, Bansal S, De Angelis D, Dye C, et al. Modeling infectious disease dynamics in the complex landscape of global health. Science. 2015;347: aaa4339. doi:10.1126/science.aaa4339

3. Metcalf CJE, Lessler J. Opportunities and challenges in modeling emerging infectious diseases. Science. 2017;357: 149–152. doi:10.1126/science.aam8335

4. Borah R, Brown AW, Capers PL, Kaiser KA. Analysis of the time and workers needed to conduct systematic reviews of medical interventions using data from the PROSPERO registry. BMJ Open. 2017;7: e012545. doi:10.1136/bmjopen-2016-012545

5. Mathes T, Klaßen P, Pieper D. Frequency of data extraction errors and methods to increase data extraction quality: a methodological review. BMC Medical Research Methodology. 2017;17: 152. doi:10.1186/s12874-017-0431-4

6. Laignelot F, Martin GL, Ossman M, Pingeon O, Boubaker A, Picovschi E, et al. Large language models show promising performance for some systematic review tasks but call for cautious implementation: a systematic review. Journal of Clinical Epidemiology. 2026;194: 112221. doi:10.1016/j.jclinepi.2026.112221

7. Li Y, Du X, Wang Y, Chen X, Zhou Z, Lian J, et al. AI-assisted literature screening: A hybrid approach using large language models and retrieval-augmented generation. International Journal of Medical Informatics. 2026;207: 106205. doi:10.1016/j.ijmedinf.2025.106205

8. Gartlehner G, Kahwati L, Hilscher R, Thomas I, Kugley S, Crotty K, et al. Data extraction for evidence synthesis using a large language model: A proof-of-concept study. Research Synthesis Methods. 2024;15: 576–589. doi:10.1002/jrsm.1710

9. Khan MA, Ayub U, Naqvi SAA, Khakwani KZR, Sipra Z bin R, Raina A, et al. Collaborative large language models for automated data extraction in living systematic reviews. Journal of the American Medical Informatics Association. 2025;32: 638–647. doi:10.1093/jamia/ocae325

10. Kwok KO, Huynh T, Wei WI, Wong SYS, Riley S, Tang A. Utilizing large language models in infectious disease transmission modelling for public health preparedness. Computational and Structural Biotechnology Journal. 2024;23: 3254–3257. doi:10.1016/j.csbj.2024.08.006

11. Fan S, Chen M, Doi SA, Ye Z, Peng Z, Tian Y, et al. Evaluating data extraction error by a large language model from randomised controlled trials: a large-scale empirical study. BMJ Evidence-Based Medicine. 2026; bmjebm-2025-114044. doi:10.1136/bmjebm-2025-114044

12. Padarha S, Kearns RO, Naidoo T, Yang L, Borchmann Ł, Błaszczyk P, et al. Evaluating AI-based Scientific Knowledge Synthesis with Epidemiological Systematic Reviews. arXiv; 2026. doi:10.48550/arXiv.2603.22327

13. WHO Collaboratory. Outbreak size. In: Ebola Outbreak Analytics CoP Resources [Internet]. 2026 [cited 1 Aug 2026]. Available: https://who-collaboratory.github.io/ebola-resources/outbreak-size/

14. Haghani M, Bliemer MCJ. Covid-19 pandemic and the unprecedented mobilisation of scholarly efforts prompted by a health crisis: Scientometric comparisons across SARS, MERS and 2019-nCoV literature. Scientometrics. 2020;125: 2695–2726. doi:10.1007/s11192-020-03706-z

15. Davidson M, Evrenoglou T, Graña C, Chaimani A, Boutron I. Comparison of effect estimates between preprints and peer-reviewed journal articles of COVID-19 trials. BMC Medical Research Methodology. 2024;24: 9. doi:10.1186/s12874-023-02136-8

16. Gwet KL. Computing inter-rater reliability and its variance in the presence of high agreement. Br J Math Stat Psychol. 2008;61: 29–48. doi:10.1348/000711006X126600

17. Lin LI-K. A Concordance Correlation Coefficient to Evaluate Reproducibility. Biometrics. 1989;45: 255–268. doi:10.2307/2532051

18. Huang J, Yang DM, Rong R, Nezafati K, Treager C, Chi Z, et al. A critical assessment of using ChatGPT for extracting structured data from clinical notes. npj Digital Medicine. 2024;7: 106. doi:10.1038/s41746-024-01079-8

19. Murton M, Boulton E, Cross S, Khan A, Kumar S, Magri G, et al. Harnessing Large-Language Models for Efficient Data Extraction in Systematic Reviews: The Role of Prompt Engineering. Cochrane Evidence Synthesis and Methods. 2025;3: e70058. doi:10.1002/cesm.70058

20. Dagdelen J, Dunn A, Lee S, Walker N, Rosen AS, Ceder G, et al. Structured information extraction from scientific text with large language models. Nature Communications. 2024;15: 1418. doi:10.1038/s41467-024-45563-x

21. Alimohamadi Y, Taghdir M, Sepandi M. Estimate of the Basic Reproduction Number for COVID-19: A Systematic Review and Meta-analysis. Journal of Preventive Medicine and Public Health. 2020;53: 151–157. doi:10.3961/jpmph.20.076

22. Park M, Cook AR, Lim JT, Sun Y, Dickens BL. A Systematic Review of COVID-19 Epidemiology Based on Current Evidence. Journal of Clinical Medicine. 2020;9: 967. doi:10.3390/jcm9040967

23. Hakki S, Zhou J, Jonnerby J, Singanayagam A, Barnett JL, Madon KJ, et al. Onset and window of SARS-CoV-2 infectiousness and temporal correlation with symptom onset: a prospective, longitudinal, community cohort study. The Lancet Respiratory Medicine. 2022;10: 1061–1073. doi:10.1016/S2213-2600(22)00226-0

24. Kasireddy E, Chow C, Collet J, Pourrahmat M-M, Fazeli MS. Evaluating the Performance of Claude 3.7 Sonnet in Data Extraction Automation for Systematic Literature Reviews. Value in Health Regional Issues. 2026;53: 101539. doi:10.1016/j.vhri.2025.101539

25. Caponio VCA, Lorenzo-Pouso AI, Magalhaes M, Ali A, Adamo D, Cirillo N, et al. Accuracy of LLMs to retrieve numeric data for meta-analysis in dentistry. Journal of Dentistry. 2026;164: 106245. doi:10.1016/j.jdent.2025.106245

26. Rokhshad R, Motie P, Shirani M, Sameie A, Revilla-León M. Performance of large language models conducting systematic review tasks in prosthodontics. Journal of Prosthetic Dentistry. 2026;136: 81–87. doi:10.1016/j.prosdent.2026.02.009

27. Li L, Mathrani A, Susnjak T. What level of automation is “good enough”? A benchmark of large language models for meta-analysis data extraction. Research Synthesis Methods. 2026;17: 671–692. doi:10.1017/rsm.2025.10066

28. Ferguson N, Pennington J, Beghian N, Mohan A, Kiela D, Agrawal S, et al. ExtractBench: A Benchmark and Evaluation Methodology for Complex Structured Extraction. arXiv; 2026. doi:10.48550/arXiv.2602.12247

29. Sujau M, Wada M, Vallée E, Hillis N, Sušnjak T. Accelerating Disease Model Parameter Extraction: An LLM-Based Ranking Approach to Select Initial Studies for Literature Review Automation. Machine Learning and Knowledge Extraction. 2025;7: 28. doi:10.3390/make7020028

30. Garcia GL, Manesco JRR, Paiola PH, Miranda L, de Salvo MP, Papa JP. A Review on Scientific Knowledge Extraction using Large Language Models in Biomedical Sciences. arXiv; 2024. doi:10.48550/arXiv.2412.03531

31. Serdiukov A, Dragvelis V, Smutin D, Taldaev A, Muravyov S. Efficient and Verified Extraction of the Research Data Using LLM. Preprints.org; 2025. doi:10.20944/preprints202511.2140.v1

32. Dao N, Quesada L, Hassan SM, Iturrioz Campo M, Johnson S, Ghose S, et al. Generative artificial intelligence for automated data extraction from unstructured medical text. JAMIA Open. 2025;8: ooaf097. doi:10.1093/jamiaopen/ooaf097

