## Supporting Information for "Using large language models to facilitate literature review and data extraction for infectious disease models: COVID-19 as a test case"

### S1 Appendix. Search strategy and screening protocol

#### *Section 1. Search Terms*

PubMed Search Terms:

(
("COVID-19"[Mesh] OR "SARS-CoV-2"[Mesh] OR "Betacoronavirus"[Mesh]) OR
("COVID-19"[Title/Abstract] OR "COVID19"[Title/Abstract] OR "COVID 19"[Title/Abstract] OR
"SARS-CoV-2"[Title/Abstract] OR "SARS-CoV2"[Title/Abstract] OR "SARSCoV2"[Title/Abstract] OR
"2019-nCoV"[Title/Abstract] OR "2019 nCoV"[Title/Abstract] OR "nCoV"[Title/Abstract] OR
"novel coronavirus"[Title/Abstract] OR "new coronavirus"[Title/Abstract] OR
"Coronavirus Disease 2019"[Title/Abstract] OR "severe acute respiratory syndrome coronavirus 2"[Title/Abstract])
)
AND
(
("Basic Reproduction Number"[Mesh] OR "Disease Transmission, Infectious"[Mesh]) OR
("R0"[Title/Abstract] OR "R 0"[Title/Abstract] OR "basic reproduction number"[Title/Abstract] OR
"basic reproductive number"[Title/Abstract] OR "basic reproductive rate"[Title/Abstract] OR
"reproductive number"[Title/Abstract] OR "reproduction number"[Title/Abstract] OR
"reproductive rate"[Title/Abstract] OR "reproduction rate"[Title/Abstract] OR
"transmission rate"[Title/Abstract] OR "transmission dynamics"[Title/Abstract] OR
"transmission parameter"[Title/Abstract] OR "transmission characteristic"[Title/Abstract] OR
"serial interval"[Title/Abstract] OR "generation time"[Title/Abstract] OR "generation interval"[Title/Abstract] OR
"incubation period"[Title/Abstract] OR "latent period"[Title/Abstract] OR "latency period"[Title/Abstract] OR
"epidemiological parameter"[Title/Abstract] OR "epidemiologic parameter"[Title/Abstract] OR
"transmissibility"[Title/Abstract] OR "force of infection"[Title/Abstract] OR
"contact rate"[Title/Abstract] OR "secondary attack rate"[Title/Abstract] OR
"effective reproduction number"[Title/Abstract] OR "Rt"[Title/Abstract] OR "R t"[Title/Abstract] OR
"doubling time"[Title/Abstract] OR "growth rate"[Title/Abstract])
)

AND
("2019/12/31"[epdat] : "2020/4/30"[epdat])

Run in the PubMed web interface on 4 January 2026, returning 750 records, which were exported page by page into a Zotero library.

medRxiv Search:

```
covid_terms <- c(
  "COVID-19", "COVID19", "COVID 19",
  "SARS-CoV-2", "SARS-CoV2", "SARSCoV2", "SARS CoV 2",
  "2019-nCoV", "2019 nCoV", "nCoV 2019",
  "novel coronavirus", "new coronavirus",
  "coronavirus 2019", "coronavirus disease",
  "Coronavirus Disease 2019",
  "severe acute respiratory syndrome coronavirus 2"
)


epi_terms <- c(
  "R0", "R 0", "R<sub>0</sub>", "R₀",
  "basic reproduction number", "basic reproductive number",
  "basic reproductive rate", "basic reproduction rate",
  "effective reproduction number", "effective reproductive number",
  "time-varying reproduction number", "reproduction number",
  "reproductive number", "reproduction rate", "reproductive rate",
  "transmission rate", "transmission probability",
  "infection rate", "attack rate", "secondary attack rate",
  "serial interval", "generation time", "generation interval",
  "incubation period", "latent period", "latency period",
  "transmission dynamics", "transmission parameter",
  "transmission characteristic", "transmissibility",
  "transmission potential",
 "doubling time", "growth rate", "contact rate", "contact pattern",
"epidemiological parameter", "epidemiologic parameter",
  "epidemic parameter", "outbreak parameter")
```

#### *Section 2. Inclusion / Exclusion Criteria for Screening*

INCLUSION CRITERIA (Must meet Condition 1 AND at least one term from Condition 2):

1. Primary research or secondary data analysis about SARS-CoV-2 / COVID-19.
2. Reports or clearly aims to estimate at least one of the following:
   a. Reproduction number (R0, Rt, Re, basic/net/effective/controlled).
   b. Serial interval (SI).
   c. Incubation period (IP).

EXCLUSION CRITERIA (Select if ANY apply):

1. Animal-only studies.
2. Letters, commentaries, editorials, response to editors (no original data/analysis).
3. Pure Case reports (clinical description only, no population-level parameter estimation).
4. Modeling papers that do NOT report or calibrate to R0/SI/IP.
5. Articles that do not provide any concrete estimate (numbers) for the parameters.
6. Not COVID-19 related / Review articles without new estimates.

#### Section 3. Search retrieval and gold standard procedure

Retrieval and deduplication. The PubMed search was run in the web interface on 4 January 2026 and its 750 records were exported page by page into a Zotero library, from which the metadata were written out as CSV. The medRxiv search was run on 9 January 2026 by an R script against the medRxiv API, which also downloaded the PDFs. Records were de-duplicated across the two sources, and where a preprint had several versions only the latest version posted on or before 30 April 2020 was retained. The metadata carried forward were the title, abstract, authors and DOI, formatted uniformly into CSV files so that the screening pipeline could read them without further preparation. Section 1 gives the full search string for each source; the medRxiv retrieval script accompanies the code release.

Gold-standard extraction protocol. The 50 articles were randomly sampled from eligible records after human screening to provide a feasible sample reflecting the range of articles and extraction complexity that would be encountered in real-world epidemiological literature. The two reviewers worked to a written protocol that specified the target fields, the uncertainty formats to be recorded, and the decision rules for handling articles that report several values for one parameter, values stratified by subgroup or setting, and values the article assumes rather than estimates. The protocol was fixed before extraction began and neither reviewer saw any model output while applying it.

Inter-rater statistics. Reportability agreement is reported with Gwet's AC1 rather than Cohen's κ because most entries are not reported, and κ is unstable when one category dominates the margins. Agreement on the numeric value is reported with Lin's concordance correlation coefficient, which is computed only on the entries both reviewers populated. The concordance coefficient measures how faithfully a recorded number is read and cannot register a disagreement about whether a value exists at all, which is what the reportability figure captures.

#### Section 4. Modified PRISMA: flow of records through the review

Fig A gives the flow of records through the human gold-standard review, following the PRISMA 2020 template for reviews that search databases only. Fig B gives the corresponding flow for the automated part. It departs from the template because a single pipeline was run independently by four models over the same records, and is labelled a modified PRISMA flow for that reason.

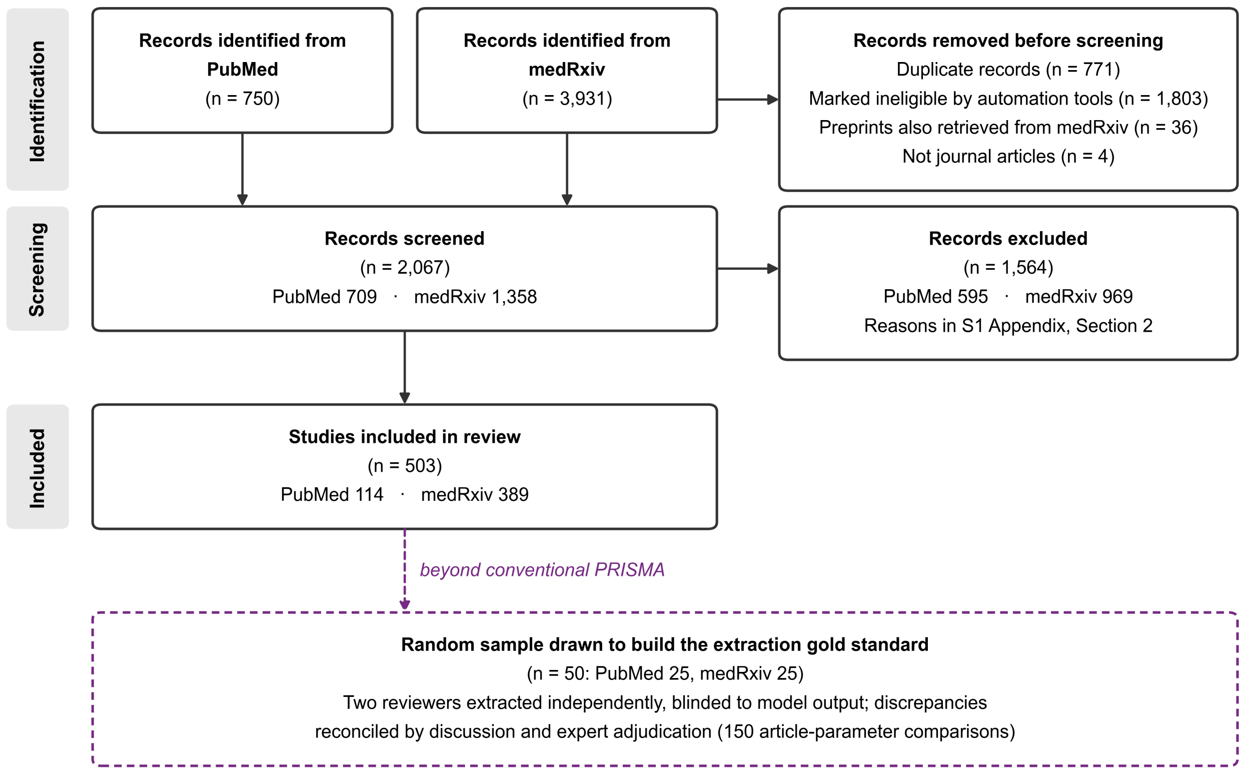


**Fig A. Flow of records through the human gold-standard review.** Solid boxes follow the PRISMA 2020 template for reviews that search databases only. Records were screened on title and abstract, with full text consulted where the decision was uncertain. The dashed box is an extension beyond the template: PRISMA ends at the included set, and this sample exists to build the gold standard against which the four language models were scored. Counts for the automated arm are given in Fig B.

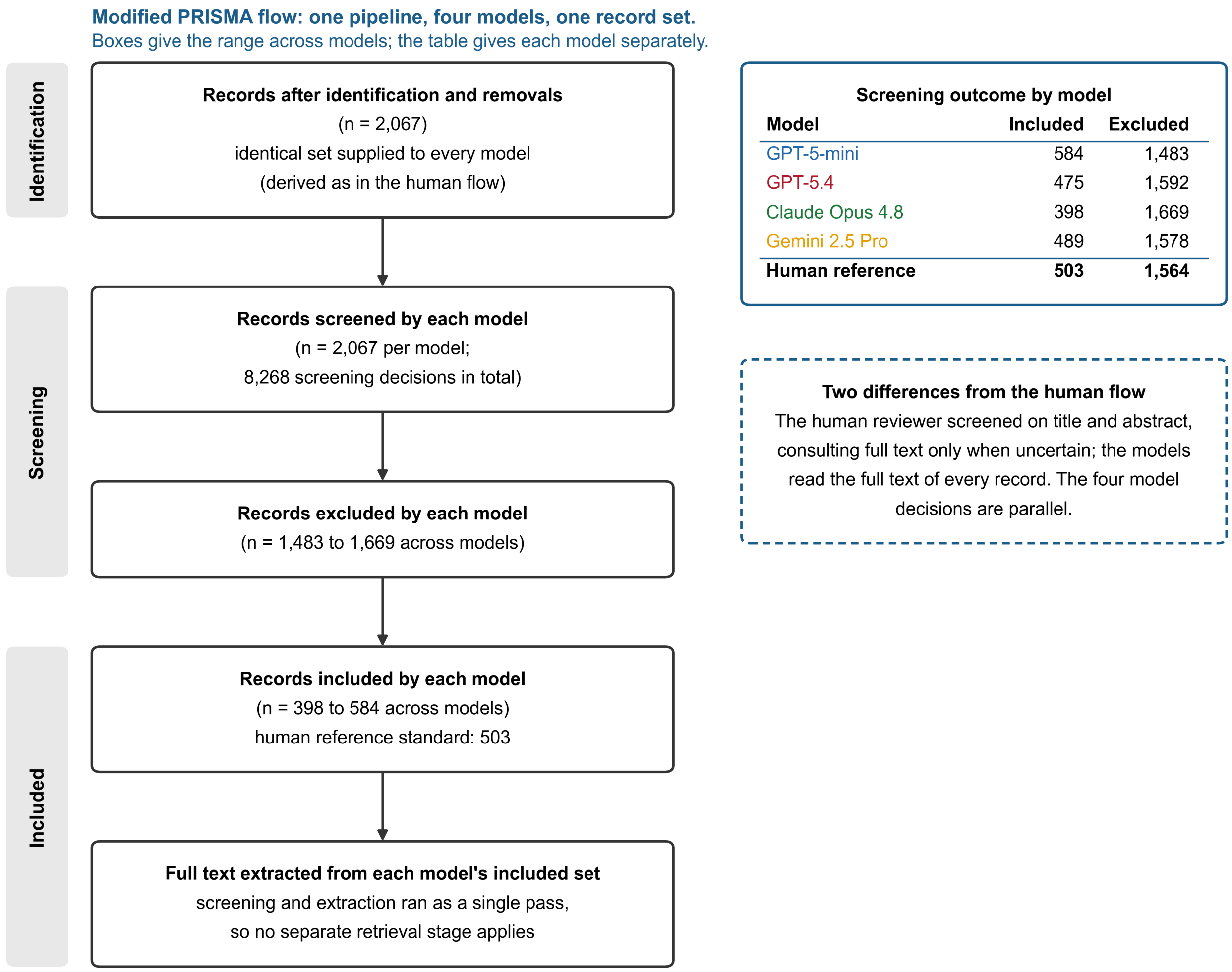


| Model | Included | Excluded |
|---|---|---|
| GPT-5-mini | 584 | 1,483 |
| GPT-5.4 | 475 | 1,592 |
| Claude Opus 4.8 | 398 | 1,669 |
| Gemini 2.5 Pro | 489 | 1,578 |
| **Human reference** | **503** | **1,564** |

**Fig B. Modified PRISMA flow for the automated arm.** A single locked pipeline was run independently by four language models over the same 2,067 records, so each box gives the range across models and the table gives each model separately. The template describes one review making one decision per record; the four decisions here are parallel, and the single column is a presentational choice. The human reviewer screened on title and abstract and consulted full text only where uncertain, whereas the models goes directly into the full text of every record. Agreement between the two arms is reported in Fig 2 of the main text.

### S2 Appendix. Pipeline implementation and prompts

#### *Section 1. Pipeline implementation and model configuration*

*Document ingestion.* To overcome the parsing failures that bottleneck out-of-the-box LLM tools, text was extracted locally with a layered strategy. Each PDF was first parsed with pdfminer to extract raw text and structural metadata; if no text was recovered, PyPDF2 was used as a fallback. Pages yielding little or no machine-readable text were rendered to images at 300 dpi and re-extracted using Tesseract optical character recognition. The recovered text was normalized to Unicode NFKC, and each page was prefixed with a positional tag of the form [p.X-Section], where the section label (Abstract, Methods, Results, etc.) was inferred from page headings. These tags preserve provenance and allow every extracted value to be traced to a specific page and section of the source, for those values whose evidence quotation is retained.

*Screening.* For each article the first 60,000 characters of tagged main text were submitted to the language model with a system prompt encoding the inclusion and exclusion criteria (Methods, Human gold standard). Text beyond that window was not seen at screening, and articles exceeding it were not re-screened at full length. The model returned a structured JSON object containing a decision (include, uncertain, or exclude), an integer certainty score (1-10), the satisfied or violated criterion, and a brief justification. Articles judged excluded were recorded and dropped, while the remaining ones proceeded to extraction.

*Extraction and aggregation.* Because epidemiological articles report parameters across many sections, the full tagged text was divided into 40,000-character chunks at sentence boundaries. Each chunk was independently submitted with a strict extraction schema (Methods, LLM-facilitated pipeline) that returned, for each of the three parameters, a numerical estimate, an uncertainty estimate with its named type (e.g., 95% confidence interval (CI), 95% credible interval (CrI), standard deviation (SD)), a descriptive summary, and up to three verbatim evidence quotes carrying their [p.X-Section] tags. A schema-enforcement step then cleaned each chunk record by coercing numeric-only fields, removing instruction artifacts, and formatting the evidence quotes. Finally, the per-chunk records for an article were reconciled by a second model call into a single record: this aggregation step removed duplicates, merged multiple valid values for the same parameter, and retained the supporting quotes. The final record for each article was written to a structured dataset (CSV and JSONL) together with per-call and per-article cost logs.

*Model configuration.* Every model was called with its provider's default decoding settings. No temperature, top-p or random seed was set, so sampling followed each provider's default rather than a deterministic configuration. What constrained the form of the output was the prompt and the schema it specified rather than the sampling. The two OpenAI models were called in JSON mode and Gemini 2.5 Pro with a JSON response type, each of which guarantees only that the reply parses, while Claude Opus 4.8 was called without an output constraint and its replies were recovered from text under a ceiling of 8,000 output tokens. Calls that failed or returned unparseable JSON were retried twice, and across 19,075 successful model calls a single reply failed to parse. Articles were processed concurrently, three at a time, with at most six requests in flight. The corpus was queried under the identifiers gpt-5-mini, gpt-5.4, claude-opus-4-8 and gemini-2.5-pro; none of these aliases is pinned by its provider to a dated snapshot, so an exact re-run cannot be guaranteed to reach the same model build.

#### *Section 2. Data Extraction System and User Prompts*

SYSTEM_PROMPT_CHUNK = """You are an evidence-focused research assistant extracting data from a SINGLE text chunk of a scientific article.

STRICT RULES:
1) Extract ONLY items EXPLICITLY stated in THIS CHUNK.
2) If an item is absent, output "N/A".
3) For numeric values, keep exact precision. Do NOT convert units.
4) Uncertainty must NAME the type (e.g., "95% CI", "CrI", "Range", "sd").
5) Prefer PRIMARY analysis over secondary/sensitivity/externally-sourced values.
6) REPRODUCTION NUMBER NAMING: If paper reports "Rt" or aliases, put numeric values in Reproduction_number_estimate and explicitly include original name in summary.
7) ASSUMED VALUES:
 - If a value is merely adopted from external literature/other viruses, do NOT populate estimate/uncertainty entry; instead put "assumed: [value]" and source details into relevant *summary* and *evidence*.
 - If authors derive a value from their own data/analysis and then "assume" it for modeling, DO populate estimate/uncertainty; in *summary*, mark "assumed after derivation" and include how it was obtained.
8) When multiple valid results exist, include ALL numeric values in the numeric-only field separated by ";", and put all descriptive labels/units/regions/methods into the corresponding *summary*.
9) EVIDENCE QUOTES: Verbatim 10–40 words with [p.X-Section] tag. Include <= 3 quotes per evidence array. If no relevant info, set array to ["N/A"].
10) QUALITY/CONSISTENCY:
 - Any non-"N/A" numeric field MUST have at least one matching quote containing the SAME numeric substrings.

OUTPUT SCHEMA (JSON only):
{
"Reproduction_number_estimate": "string",
"Reproduction_number_uncertainty": "string",
"Reproduction_number_summary": "string",
"Incubation_estimate": "string",
"Incubation_uncertainty": "string",
"Incubation_summary": "string",
"Serial_interval_estimate": "string",
"Serial_interval_uncertainty": "string",
"Serial_interval_summary": "string",
"Study_design": "string",
"Geo_time": "string",
"Sample_population": "string",
"Subgroup_results": "string",

"Citation": "string",
"R0_evidence": ["string"],
"Incubation_evidence": ["string"],
"Serial_evidence": ["string"],
"Other_evidence": ["string"]
}
"""

USER_PROMPT_TEMPLATE = """Based on THIS CHUNK ONLY, extract the following items about COVID-19 transmission modeling.

**CRITICAL FORMATTING RULES FOR SUMMARIES:**
- Do NOT start summaries with phrases like "If it's R0" or "Based on...".
- Start DIRECTLY with the name and value.
- Example for R0: "R0: 2.5 (95% CI 2.0-3.0) estimated using EG method..."
- Example for Rt: "Rt: 1.2 (CrI 1.0-1.4) for London region..."
- Example for Serial Interval: "5.2 days (sd 2.8) based on 100 pairs..."

Return JSON with EXACTLY these keys:
{
  "Reproduction_number_estimate": "string",      // numbers only, connect multiple values with ';', "N/A" if value is not original and "assumed"
  "Reproduction_number_uncertainty": "string",      // E.g., "95% CI 2.1–2.8; CrI 1.2–2.0", "N/A" if value is not original and "assumed"
  "Reproduction_number_summary": "string",      // START DIRECTLY with label/value. E.g. "R0: 2.5..." or "Rt: 1.1...".
  "Incubation_estimate": "string",      // numbers only, connect multiple values with ';', "N/A" if value is not original and "assumed"
  "Incubation_uncertainty": "string",      // "N/A" if value is not original and "assumed"
  "Incubation_summary": "string",      // E.g. "5.2 days (95% CI...)", "Assumed 3.4 days (95% CI 2.8-4.0) based on [p.X-Section]..."
  "Serial_interval_estimate": "string",      // numbers only, connect multiple values with ';', "N/A" if value is not original and "assumed"
  "Serial_interval_uncertainty": "string",      // E.g. "sd 2.0; 95% CI 1.0-3.0", "N/A" if value is not original and "assumed"
  "Serial_interval_summary": "string",      // E.g. "5.2 days (sd 2.8)", "Assumed 4.0 days (95% CI 3.0-5.0) based on [p.X-Section]..."
  "Study_design": "string",
  "Geo_time": "string",
  "Sample_population": "string",
  "Subgroup_results": "string",
  "Citation": "string",
  "R0_evidence": ["string","string","string"],
  "Incubation_evidence": ["string","string","string"],

```
  "Serial_evidence": ["string","string","string"],
  "Other_evidence": ["string","string","string"]
}
Text chunk:
---
{chunk_text}
---
"""
SYSTEM_PROMPT_AGG = """You are reconciling multiple JSON extractions from DIFFERENT CHUNKS of the SAME article into ONE final JSON using the SAME schema.

RESOLUTION:
1) Consider ONLY candidates whose non-"N/A" numeric fields are supported by quotes containing the same numeric substrings.
2) Prefer primary analysis in main text; prefer explicit uncertainty with type+bounds.
3) When multiple valid results exist, MERGE numeric values by semicolon in numeric-only fields; combine narrative details for each value in corresponding summary.
4) Keep units/descriptors/labels in summaries; keep numeric-only fields strictly numeric (with uncertainty types allowed in *_uncertainty).
5) Merge evidence arrays and keep <= 3 quotes per array; preserve the "[p.X-Section]" tags and any figure/table wording.
6) If nothing valid remains, use "N/A" and ["N/A"] for its evidence array.
OUTPUT: strictly the same JSON schema as chunk-level.
"""
USER_PROMPT_AGG_TEMPLATE = """Reconcile following JSON candidates.
JSON candidates:
---
{candidates_json}
---
"""
```

### S3 Appendix. Error taxonomy, observed patterns and representative cases

#### *Section 1. Error category definitions*

1. **Identity confusion and provenance confusion:** The model reads a value correctly but files it under the wrong parameter, or treats a borrowed value as the study's own. These are two distinct failures that often co-occur in the same article, and we treat them together because they share a remedy. Identity confusion returns a neighbouring quantity rather than the one requested (a generation time returned as a serial interval, a latent period as an incubation period, an epidemic growth rate as a reproduction number). Provenance confusion returns the right parameter, but not the study's own primary estimate (an assumed model input, a value cited from another study, or an estimate for another pathogen such as SARS-CoV-1 or influenza).
2. **Granularity Omission:** When a study reports many stratified estimates (e.g., region- or cohort-specific values), the model returns a correct representative summary, such as the overall estimate with its minimum and maximum, rather than transcribing every value.
3. **Screening Exclusion:** A relevant, human-included article that the model rejects at screening. Because the article never reaches extraction, every value it reports is a false negative. We account for it separately as a screening-cascade effect (Results, Full-text data extraction accuracy) and report a scope restricted to the human-included articles.

#### *Section 2. Observed error patterns by category*

Across the eight model–corpus evaluations, 320 gold-standard values were assessed independently by four models, giving 1,280 model–gold-value comparisons. Of the 649 false-negative value comparisons, 536 (82.6%) were large-granularity omissions, 65 (10.0%) arose because the article had been excluded at screening, and 48 (7.4%) were other in-article omissions. The models returned 653 values, of which 22 (3.4%) were false positives attributable to identity or provenance confusion. All the 22 cases have values present in the source articles; none was fabricated. A gold value missed by more than one model contributes once for each model, consistent with the model-specific results in Fig 4.

Under identity confusion, the models conflated distinct but neighbouring epidemiological quantities, such as placing a printed generation time in the serial interval field, including in studies that estimated no serial interval at all (Table B).

Similarly, some LLMs extracted latent periods in the incubation period field, and the epidemic growth rate in the reproduction number field. However, even when a model misassigned a value it usually named the quantity correctly in its output: in eight of the nine misassigned cases the model summary either identified the true quantity or flagged the value as assumed or cited. The models also identified parameters from a mathematical definition where the name never appeared. One read an incubation period from a "mean interval from contact to onset of symptoms," and another inferred a five-day incubation period from an assumed transition rate of one-fifth per day stated only in a supplementary table.

Under provenance confusion, models placed assumed or external values in primary fields while flagging the provenance in their summaries: an R0 of 2.28 recorded as "assumed … as a model parameter" {Chatterjee, 2020}, an incubation of 5 days as "cited from external source [5], not derived in this study" {Sebastiani, 2020}, and an R0 of 1.92 as pertaining to "JpnInf2019" (medRxiv 2020.02.26).

For granularity omission, the models tend to summarize findings rather than enumerate high-granularity reporting, often retaining a representative estimate and range from papers that tabulate a parameter across dozens of strata, such as a national R0 with its minimum and maximum drawn from values across 50 states. This pattern reduced value-level recall. In the sensitivity analysis, the alternative output configuration recovered substantially more values, although the compared configurations also differed in reconciliation instructions and limits on retained quotations (S4 Appendix).

A hallucination was defined as a numeric value with no counterpart anywhere in the source article. No such hallucinations were observed in this study.

**Table A. Representative extraction errors. The table includes identity or provenance errors and granularity omissions. In every case, the value was present in the source text; none was fabricated. The examples include all four models.**

| # | Model(s) that produced the error | Corpus · Article | Parameter | Gold value | Model output | What the value actually was (detail) | Source location (section · table/figure) | Cause of error (from article context) |
|---|---|---|---|---|---|---|---|---|
| 1 | GPT-5-mini | medRxiv · Kermack–McKendrick model (2020.02.26) | R0 | N/A (no COVID R0) | "R0: 1.92 estimated … for JpnInf2019 using the Kermack–McKendrick model" | Japanese-influenza R0 (the paper's validation example), not a COVID-19 estimate. | Results/Discussion prose — "relations are applied to influenza in Japan in 2019"; the value is computed from the influenza-fit parameters stated in text (not in a table) | The article develops a method for COVID-19 but reports this R0 for an influenza validation example. The output assigned the value according to the article-level topic rather than the pathogen attached to the estimate. |
| 2 | Gemini 2.5 Pro, GPT-5-mini | PubMed · Fang et al. (intervention model) | Incubation | N/A | "Assumed average latency (Te) of 7 days for the SEIR model simulation" (GPT-5-mini similar) | An assumed model latency Te = 7 d, not an estimated incubation period. | Methods — SEIR model parameterization (assumed Te = 7 d) | Gemini 2.5 Pro and GPT-5-mini identified the value as an assumed latency (Te) but placed it in the incubation field. GPT-5.4 returned N/A. |

| | | | | | | | | |
|---|---|---|---|---|---|---|---|---|
| 3 | GPT-5-mini | medRxiv · Wuhan travellers (Backer; 2020.01.27) | Incubation | Weibull: 6.4 (95% CrI: 5.6-7.7); Gamma: 6.5 (95% CrI: 5.6-7.9); Lognormal: 6.8 (95% CrI: 5.7-8.8) | "Incubation period: 6.4 days (95% credible interval 5.6 – 7.7) mean using Weibull fit… based on 88 travellers from Wuhan. [p.6-Results]" | Gamma-fit mean 6.5 (95% CI 5.6–7.9) and Lognormal-fit mean 6.8 (95% CI 5.7–8.8) missed; only the Weibull 6.4 kept. | Results table (distribution-fit table: Weibull/Gamma/Lognormal means with 95% CI), also noted in Results text | The output retained the Weibull estimate but omitted the Gamma and Lognormal alternatives reported in the same table. |
| 4 | GPT-5-mini, Gemini 2.5 Pro | PubMed · Chen et al. (136 cases) | Incubation | All cases (N=136): 7.7 ± 4.1, imported cases (N=29) 6.7 ± 4.1, and local cases (N=107) 8.4 ± 4.1 days. | "The mean incubation period for all 136 cases was 7.7 days (sd 4.1) …" (GPT-5-mini similar) | Imported-case mean 6.7 d and local-case mean 8.4 d (both ±4.1) missed; only the overall 7.7 d kept. | Table 1 ("Epidemiological and clinical characteristics … imported cases, local cases, second-generation cases") — the subgroup incubation means are table rows; 7.7 overall is in the abstract/text | Both outputs retained the overall mean and omitted the imported-case and local-case subgroup means reported in Table 1. |
| 5 | All four (GPT-5.4, GPT-5-mini, Gemini 2.5 Pro, Claude Opus 4.8) | medRxiv · 50-state testing model (2020.04.20) | R0 | 104 state-level values | "R0 for the US … 3.45 …; state-level R0 ranged from 1.92 … (South Dakota) to 5.17 … (Missouri) …" (GPT-5.4 shown; all four similar) | Per-state R0 for 51 states × 2 methods (104 values) not enumerated beyond the national mean and min/max. | Supplementary Table S2 (per-state R0, both case- and test-based); national 3.45 and the min/max are stated in the Results text | All four outputs retained the national mean and range but did not enumerate the per-state values in Supplementary Table S2. |
| 6 | Gemini 2.5 Pro | medRxiv · Italy regional R0/Rt (ISS; 2020.04.08) | R0 / Rt | Six regional R0 (Lombardia 2.96 … Puglia 2.61); range 2.13–3.33; Rt max ≈ 3 (week of Feb 17–23); Rt ≈ 2.5–3 (until Mar 4–8) | "R0: 2.96 … Lombardia; … 2.61 … Puglia. The net reproduction number (Rt) showed a decreasing trend in northern regions." | The two quantitative Rt-trend values (≈3; 2.5–3) reported only in narrative, omitted as numbers. | Results prose (Rt-estimation section) — "...reaching maximum values around 3 over the week from February 17 to 23"; "...nearly constant at values around 2·5-3 until March 4-8" (not in a table; the six regional point estimates are in a table, which Gemini captured) | Gemini 2.5 Pro retained the regional point estimates but omitted two approximate Rt values reported only in prose. GPT-5.4, GPT-5-mini, and Claude Opus 4.8 captured both values. |
| 7 | Gemini 2.5 Pro, Claude Opus 4.8 | PubMed · Liu et al. (pre-symptomatic transmission) | Incubation | 6.0 (95% eCI 1.3–17.2) Bi base; 5.2 (1.1–14.3) Li; 6.0 (1.6–14.0) Bi | "6.0 days (95% eCI 1.3–17.2) … from Bi et al. … 5.2 days (1.1–14.3) … adopted from external sources." (Claude shown; Gemini similar) | Third incubation value 6.0 d (95% eCI 1.6–14.0), used in a combined scenario, missed. | Table 1 ("Overview of scenarios tested …") — the 6.0 (1.6–14.0) appears only as a scenario row in Table 1; the base-case 6.0 (1.3–17.2) and the 5.2 (1.1–14.3) are stated in the Results prose | Both outputs retained the two incubation values described in prose but omitted the third near-duplicate value reported only in the scenario table. GPT-5.4 and GPT-5-mini captured all three values. |

**Table B. Identity and provenance labelling in the summary field. Representative cases, drawn from all four models, in which the output named the returned quantity or marked it as assumed, external, or absent (Results, Error classification and root cause analysis).**

| Model | Corpus · Article | Requested field | Verbatim summary (self-label / provenance flag) | What the labelling preserves |
|---|---|---|---|---|
| GPT-5.4 | PubMed · Matrajt & Leung | Incubation | "Mean latent period 5.16 days (range 4.5–5.8 days) … the model considered the latent period equal to the incubation period." | Identifies the quantity as a latent period and states the authors' assumption that it equals the incubation period. |
| GPT-5.4 | PubMed · Shim et al. | Serial interval | "No original serial interval estimate was reported. The analysis assumed a generation interval … mean 4.41 days …" | Flags the requested parameter as absent and names the assumed surrogate used in its place. |
| Claude Opus 4.8 | PubMed · Fang et al. | Serial interval | "Generation period Tg (mean 8.4 days), approximate to serial interval … adopted from external source, not originally estimated." | Distinguishes generation time from serial interval and marks it external and not original; GPT-5.4 reached the same distinction. |
| Gemini 2.5 Pro | PubMed · Sebastiani et al. | Incubation | "Assumed a 5-day median interval between infection and symptoms based on external source [5] Lauer et al. … a priori information …" | Marks the value as an external, a priori input rather than a quantity estimated in the study. |
| Gemini 2.5 Pro | PubMed · Fang et al. | Incubation | "Assumed average latency (Te) of 7 days for the SEIR model simulation." | Names an assumed model latency, not an estimated incubation period. |
| GPT-5-mini | medRxiv · transportation-network model | Serial interval | "Generation time 8.4 days …; generation time 7 days …" | Identifies the quantity as a generation time, although the value remains in the serial-interval field. |

#### *Section 3. Screening losses due to varied human vs. AI approaches*

Across the four models, 295 relevant articles were wrongly excluded: 131 by Claude Opus 4.8, 75 by Gemini 2.5 Pro, 62 by GPT-5.4, and 27 by GPT-5-mini. Of these decisions, 217 were recorded under exclusion criterion 4, which removes modelling papers that do not report or calibrate to a target parameter. The human gold standard retained articles that used a published parameter as a model input, whereas criterion 4 excluded such articles when they did not also report or calibrate to a target parameter. In 136 of the 217 decisions, the explanation used at least one of the words assumed, adopted, cited, or borrowed. The remaining 78 misses were recorded under criterion 5 (47), criterion 6 (17), or criteria 2 and 3 (14); Table C gives examples. This distribution indicates a mismatch between the screening criteria and gold-standard eligibility. Screening false positives occurred most often for GPT-5-mini and Gemini 2.5 Pro.

**Table C. Representative screening errors (false negatives and false positives) and their causes, traced to the source article and the four models' screening output (decision, certainty, criteria term).**

| # | Dir. | Model(s) with the wrong call | Corpus · Article | Human gold (includable parameter) | AI decision · certainty · criteria_term | AI screening_explanation (quoted) | What the article actually reports & where | Cause of the mis-screen |
|---|---|---|---|---|---|---|---|---|

| | | | | | | | | |
|---|---|---|---|---|---|---|---|---|
| 1 | FN | Claude only (GPT-5.4, mini, Gemini included) | PubMed · Chen et al. (136 cases, Chongqing ) | Include — incubation 7.7 d (+ subgroups) | exclude · c7 · t5 | "…time from contact symptomatic case to illness, which is called the incubation period, was 7.7 days" which is a descriptive average rather than a formal incubation period distribution estimation | Incubation 7.7 d (Results text/abstract) + subgroup means in Table 1 | The explanation treated the computed mean as descriptive because the article did not report a fitted distribution. The other three models included the article based on its incubation-period estimate. |
| 2 | FN | Claude only (GPT-5.4, mini, Gemini included) | PubMed · Jin et al. (74 cases) | Include — incubation median 4 d (IQR 3–7) / 5 d | exclude · c8 · t5 | "…While it reports a calculated incubation period (median 4 … 3–7 …), the R0 values mentioned (2.26 and 2.6…" [are cited] | Incubation median 4 d (IQR 3–7) for GI; 5 d non-GI (Results) | The explanation focused on the cited R0 values and did not retain the article based on its calculated incubation periods. |
| 3 | FN | GPT-5.4 only (mini, Gemini, Claude included) | PubMed · Torres-Roman et al. (Peru R0, "Dear Editor" letter) | Include — R0 2.97 (Peru) / 2.88 (Lima) | exclude · c9 · t2 | "explicitly presented as a 'Dear Editor' letter. Although it contains an original estimate of the basic reproduction number (R0=2.97 for Peru; 2.88 for Lima), the review excludes letters/commentaries/editorials even when they include analysis" | Original R0 = 2.97 (Peru) / 2.88 (Lima) estimated from early case counts; stated in a "Dear Editor" letter (text) | The decision applied exclusion criterion 2 based on publication type despite identifying original R0 estimates in the article. The other three models included the article based on those estimates. |
| 4 | FN | GPT-5.4 only (mini, Gemini, Claude included) | PubMed · Muniz-Rodriguez et al. (Transmission potential, Iran 2020; research letter) | Include — R0 4.4, Re 3.5→1.55; serial interval 4.41 d | exclude · c10 · t 2 | "labeled as a research letter. Although it contains original COVID-19 analysis and reports concrete reproduction number estimates (R0 and Re), the review excludes letters/research letters" | R0 ≈ 4.4 and time-varying Re (3.5→1.55) plus serial interval 4.41 d, estimated for Iran's early epidemic (Methods/Results of the research letter) | The decision applied exclusion criterion 2 because the article was labelled a research letter, despite identifying original R0 and Re estimates and a serial interval in the explanation. |

| | | | | | | | | |
|---|---|---|---|---|---|---|---|---|
| 5 | FN | GPT-5.4 only (mini, Gemini, Claude included) | medRxiv · 34 children, West China (2020.03.12) | Include — incubation median 10.5 d (IQR 7.75–25.25) | exclude · c10 · t 3 | "a COVID-19 paediatric multiple-center case series … Although it reports a median incubation period of 10.50 (7.75–25.25) days … the incubation period is presented descriptively within a case series rather than as a dedicated population-level transmission parameter analysis" | Median incubation 10.5 d (IQR 7.75–25.25) computed for 34 paediatric cases (Results/Table) | The decision applied criterion 3 because the article was a case series and required a dedicated population-level analysis, a condition not specified in the screening protocol. The other three models included the article based on its calculated median incubation period. |
| 6 | FN | GPT-5.4, Gemini (mini, Claude included) | PubMed · Nishiura et al. (Wuhan initial cluster; J Clin Med, labeled "Editorial") | Include — serial interval mean 7.4 d; generation-dependent R (3.0/1.3/6.7/0.2) | exclude · c10 · t 2 | GPT-5.4: "labeled as an editorial. Although it reanalyzes early COVID-19 data and reports concrete estimates for serial interval and generation-dependent reproduction numbers, the exclusion criteria state that … editorials should be excluded"; Gemini: "labeled as an 'Editorial' on page 1, which falls under the exclusion criteria for article type" | A reanalysis of the initial Wuhan cluster estimating SI mean 7.4 d and generation-dependent R; §2 "Epidemiological Analysis." The journal's type label is "Editorial" | Both decisions applied criterion 2 because the article was labelled an Editorial, despite its original serial-interval and reproduction-number estimates. GPT-5.4 identified the estimates in its explanation, whereas Gemini 2.5 Pro cited the page-one publication label. |
| 7 | FP | GPT-5.4, Gemini (mini, Claude excluded) | medRxiv · Gray et al. ("No test is better than a bad test," 2020.04.16) | Exclude — testing-strategy SIR model; R0 only as a definitional formula | include · c9 · t a | GPT-5.4: "reports/calculates the basic reproduction number in the appendix, giving R0 = beta/gamma … beta = 0.32 and gamma = 0.1, implying R0 = 3.2"; Gemini: "The appendix defines R0 = β/γ and provides a sample calculation (R0=3) … Table 2 lists … (β=0.32, γ=0.1)" | A modified SIR model on diagnostic uncertainty in mass testing; the only R0 content is the textbook identity R0 = β/γ with assumed β=0.32, γ=0.1 (Appendix / Table 2) — no R0 estimated from data | Both decisions treated R0 calculated from assumed β and γ values as an article-specific estimate. GPT-5-mini and Claude Opus 4.8 excluded the article under criterion 4. |

| | | | | | | | | |
|---|---|---|---|---|---|---|---|---|
| 8 | FP | GPT-5-mini only (GPT-5.4, Gemini, Claude excluded) | PubMed · Chawla et al. (perinatal-neonatal management guideline) | Exclude — clinical-practice guideline; no original estimate | include · c9 · t a,c | “The paper states ‘The average number of people infected by one infected individual is between two to three,’ which reports an estimate of the reproduction number. … ‘Incubation period varies from 2-14 days with a median of 5 days’” | A FOGSI management guideline; the “two to three” line and “2–14 d, median 5” are generic background restated from the literature (introductory text), not values the document derives | The decision attributed two generic background values to the guideline itself. The other three models excluded the article based on publication type or the absence of original estimates. |
| 9 | FP | GPT-5-mini, Gemini (GPT-5.4, Claude excluded) | PubMed · Yong et al. (serological cluster-linking; Lancet Infect Dis) | Exclude — cluster-linking serology study; no population-level estimate | include · c9–10 · t c | GPT-5-mini: “reports incubation period data (Figure 2 shows incubation periods … for 30 cases)”; Gemini: “reports on the incubation period for 30 individuals … visualized in Figure 2, titled ‘Incubation period, duration of symptoms, and length of admission’” | A serological investigation linking three Singapore clusters; Figure 2 draws each of 30 patients’ timelines (an incubation bar per case) but the paper computes no summary incubation statistic — both models extracted no numeric value | The decisions treated patient-level incubation bars as an includable parameter estimate, although the article reported no population-level summary and both extraction outputs contained no numeric value. GPT-5.4 and Claude Opus 4.8 excluded the article under criterion 5. |

### S4 Appendix. Sensitivity analysis: output format, input mode and aggregation

#### Section 1. Output format, input mode and aggregation on dense-target sensitivity set

This sensitivity analysis used a deliberately dense subset rather than the 50-article gold standard as a whole. Five article-parameter pairs from the four gold-standard articles with the highest value counts contained 12, 15, 15, 18, and 104 values; the 104-value article therefore supplied 104 of the 164 values in each repetition. GPT-5.4 was used for three repetitions of every cell, giving 492 scored values per row. The design is summarized in Methods, Sensitivity analysis.

Four elements were varied. The output format stored all values for a parameter in one field, as in the deployed pipeline, or used one row per reported value. The instruction used either the original locked wording or a variant requiring every individual value and prohibiting compression. Input consisted of locally extracted text or page images. Repeated candidates were reconciled by the model or combined by a fixed rule that retained every distinct value. The last three factors were crossed under the row format in a 2 × 2 × 2 factorial design, with both single-field instruction arms evaluated alongside it. A separate small-granularity control set compared the baseline and alternative configurations on articles with few reported values.

**Table A. Output format, input mode, and aggregation on the dense-target sensitivity set. GPT-5.4 was evaluated on five article-parameter targets. Each row contains 492 scored values from 15 extraction jobs, all of which were scorable. Recall and precision are value-level metrics for point estimates. Single-field configurations used model reconciliation because fixed-rule aggregation applies only to row-wise candidates.**

| Output format | Input | Instruction | Aggregation | TP | FP | FN | Recall | Precision |
|---|---|---|---|---|---|---|---|---|
| Single field | Extracted text | Locked | Model reconciliation | 84 | 0 | 408 | 0.171 | 1.000 |
| Single field | Extracted text | Exhaustive | Model reconciliation | 129 | 5 | 363 | 0.262 | 0.963 |
| One row per value | Extracted text | Locked | Model reconciliation | 434 | 39 | 58 | 0.882 | 0.918 |
| One row per value | Extracted text | Locked | Deterministic union | 435 | 62 | 57 | 0.884 | 0.875 |
| One row per value | Extracted text | Exhaustive | Model reconciliation | 431 | 48 | 61 | 0.876 | 0.900 |
| One row per value | Extracted text | Exhaustive | Deterministic union | 435 | 77 | 57 | 0.884 | 0.850 |
| One row per value | Page images | Locked | Model reconciliation | 234 | 17 | 258 | 0.476 | 0.932 |
| One row per value | Page images | Locked | Deterministic union | 236 | 37 | 256 | 0.480 | 0.864 |
| One row per value | Page images | Exhaustive | Model reconciliation | 433 | 34 | 59 | 0.880 | 0.927 |
| One row per value | Page images | Exhaustive | Deterministic union | 435 | 49 | 57 | 0.884 | 0.899 |

Paired contrasts were calculated as the mean difference in article-parameter recall over the 15 pairs, with 95% cluster-bootstrap intervals over the four articles. Relative to extracted text, page images changed recall by -0.065 (-0.238 to 0.000); the exhaustive instruction changed recall by +0.062 (-0.002 to 0.238); and deterministic union changed recall by +0.010 (0.000 to 0.021). The input-by-instruction interaction was +0.069 (0.000 to 0.238). This interaction was driven mainly by the 104-value article: page images under the locked wording returned 114 of 312 gold-standard values, compared with 312 of 312 for extracted text. The other four targets differed by no more than two values in any cell. All remaining two-way and three-way interactions were at most 0.004 in absolute value.

**Table B. Recall by target, pooled over three repetitions. Column headings show the number of gold-standard values per repetition. Under the locked instruction, single-field recall declined from 0.917 on the 12-value target to 0.048 on the 104-value target, while the row configuration reached 1.000 on both. Page images and extracted text differed materially only for the 104-value article under the locked wording, accounting for the input-by-instruction interaction. Targets were Wang L 2020 (medRxiv 10.1101/2020.02.29.20029421), Zhao S 2020 (Int J Infect Dis 2020;92:214-217), Pitzer VE 2020 (medRxiv 10.1101/2020.04.20.20073338), and Li M 2020 (medRxiv 10.1101/2020.04.15.20065946); the last contributed both an incubation-period and a serial-interval target.**

| Configuration | Wang L 2020, R0 (12) | Zhao S 2020, R0 (18) | Pitzer VE 2020, R0 (104) | Li M 2020, incubation (15) | Li M 2020, serial interval (15) |
|---|---|---|---|---|---|
| Single field, locked | 0.917 | 0.111 | 0.048 | 0.333 | 0.333 |
| Single field, exhaustive | 1.000 | 1.000 | 0.029 | 0.333 | 0.333 |
| Row, text, locked | 1.000 | 1.000 | 1.000 | 0.400 | 0.333 |
| Row, text, exhaustive | 1.000 | 1.000 | 1.000 | 0.400 | 0.333 |
| Row, page images, locked | 1.000 | 1.000 | 0.365 | 0.378 | 0.333 |
| Row, page images, exhaustive | 1.000 | 1.000 | 1.000 | 0.400 | 0.333 |

Across the three repetitions, recall for the locked single-field configuration was 0.177, 0.177, and 0.159; recall for the row configuration was 0.884 in each repetition. The single-field configuration returned a mean of 2 to 11 values per article across targets, whereas the row configuration returned counts closer to the gold standard, including up to 109 values for the 104-value article. For that article, the single-field candidate count was 5 in one repetition and 107 in each of the other two, but reconciliation retained 5 values in all three final records.

**Table C. Results for reserved small-granularity control articles under the baseline single-field configuration and the alternative exhaustive row configuration. Each configuration comprised nine extraction jobs and 15 gold-standard values. Both recovered all 15 values. The alternative configuration returned 24 false positives, compared with none for the baseline, corresponding to precision of 0.385 and 1.000.**

| Configuration | Gold values | TP | FP | Recall | Precision |
|---|---|---|---|---|---|
| Single field, locked, extracted text | 15 | 15 | 0 | 1.000 | 1.000 |
| Row, page images, exhaustive | 15 | 15 | 24 | 1.000 | 0.385 |

**Table D. Cross-model comparison of the baseline and alternative output configurations, with one repetition per model. Each model was run on five dense targets using the deployed single-field configuration and the row configuration with page images and exhaustive instructions. The denominator was 164 gold-standard values and five extraction jobs per cell. In the Claude Opus 4.8 row condition, one job returned an unscorable record; the remaining four targets contained 146 values.**

| Model | Output format | TP | FP | FN | Recall | Precision |
|---|---|---|---|---|---|---|
| GPT-5-mini | Single field | 44 | 3 | 120 | 0.268 | 0.936 |
| GPT-5-mini | One row per value | 124 | 49 | 40 | 0.756 | 0.717 |
| GPT-5.4 | Single field | 29 | 0 | 135 | 0.177 | 1.000 |
| GPT-5.4 | One row per value | 145 | 18 | 19 | 0.884 | 0.890 |
| Claude Opus 4.8 | Single field | 44 | 0 | 120 | 0.268 | 1.000 |
| Claude Opus 4.8 | One row per value | 127 | 17 | 19 | 0.870 | 0.882 |

| | | | | | | |
|---|---|---|---|---|---|---|
| Gemini 2.5 Pro | Single field | 28 | 0 | 136 | 0.171 | 1.000 |
| Gemini 2.5 Pro | One row per value | 125 | 54 | 39 | 0.762 | 0.698 |

#### *Section 2. Output configuration and granularity omission*

Across five dense-parameter article targets, the baseline single-field configuration achieved value-level recall of 0.171, compared with 0.884 for the alternative row configuration, using an identical denominator of 492 scored values. Under the single-field format, the exhaustive instruction increased recall to 0.262; under the row format, it produced no further increase. Table A gives the full factorial results, and Table D shows the same ordering across all four models. Because the formats also used different reconciliation instructions and quotation limits, the results support a substantial contribution from the alternative output configuration but do not isolate the effect of field structure.

Differences were larger on denser targets. The single-field format recovered 0.917 on an article with 12 reproduction numbers and 0.048 on the 104-value article, while the row format reached 1.000 on both. Single-field precision was 1.000 under the locked wording and 0.963 under the exhaustive instruction. During extraction, 288 of 492 gold values were identified, but 84 remained after reconciliation. Thus, 204 were removed during reconciliation and another 204 were absent from the segment-level candidates.

The alternative configuration mainly addressed granularity omissions and sometimes reduced precision. Results changed little for an article reporting incubation period, serial interval, and generation interval together because its remaining errors involved parameter identity. On the small-granularity controls, both configurations recovered every gold value, but the alternative configuration returned 24 false positives and the baseline returned none (Table C). This precision trade-off supports further evaluation of row-wise extraction for dense articles rather than universal use.

### S5 Appendix. Evaluation metrics and interval construction

Classification metrics. Let TP, FP, TN and FN denote the counts defined in Methods, Evaluation metrics and statistical analysis. Sensitivity, which is equivalent to recall, is the proportion of truly positive items recovered, TP/(TP+FN). Precision is the proportion of returned items that are correct, TP/(TP+FP). Specificity is the proportion of negatives correctly rejected, TN/(TN+FP). Accuracy is the overall proportion of correct decisions, (TP+TN)/(TP+TN+FN+FP).

The F-measure combines precision and recall as $F\beta = (1 + \beta^2) \times \text{precision} \times \text{recall} / (\beta^2 \times \text{precision} + \text{recall})$. F1, with $\beta = 1$, is their harmonic mean and weights the two equally. F2, with $\beta = 2$, weights recall four times as heavily as precision.

Confidence intervals. All intervals are 95%. For the proportion-based metrics, namely screening sensitivity, specificity and precision together with article-level accuracy, we used the Wilson score interval. For the area under the ROC curve we used a bootstrap over articles. For the value-level extraction metrics recall, F1 and F2 we used a cluster bootstrap that resamples articles rather than values, with 2,000 replicates and a percentile interval. Value-level precision is reported with the Wilson interval.

Model comparison. Screening decisions were compared with McNemar's test, which pairs the two models on the articles both of them saw. The value-level extraction metrics were compared with paired cluster-bootstrap tests over the same resampled articles, so that the comparison and the intervals rest on the same resampling unit.

### S6 Appendix. Transmission model and downstream comparison

#### *Section 1. Transmission model simulation*

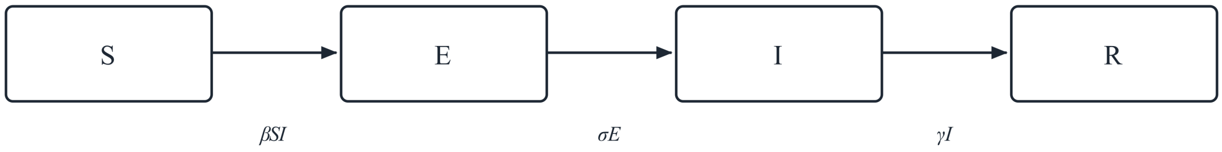


***Fig A. SEIR model diagram.***

We used an SEIR model to simulate transitions between susceptible (S), exposed (E), infected (I), and recovered (R) compartments. Susceptible individuals become infected by contacting infectious individuals at a rate, $\beta$, and move to the exposed group (E). $\beta$, defined as the per capita rate of effective contact, is calculated by dividing the basic reproduction number, R0, by the total population (N=1) and the infectious period (D=5 days). The exposed individuals develop symptoms (I) after the incubation period, at a rate $\sigma$. Finally, they recover at a rate $\gamma$ and move to the recovered group (R). The model assumes that all persons are initially susceptible, that the population size is fixed, and that mixing is homogeneous, with the initial number of infectious persons set at 1% of the population. The following differential equations describe the transmission dynamics.

**Table A. Differential equations for the SEIR transmission dynamic model**

| | |
|---|---|
| $\frac{dS}{dt} = -\beta SI$ | where<br>$S + E + I + R = N$<br>Susceptible + Exposed + Infected + Recovered = N |
| $\frac{dE}{dt} = +\beta SI - \sigma E$ | $N =$ Population<br>$\beta = R_0\gamma$ |
| $\frac{dI}{dt} = +\sigma E - \gamma I$ | $\sigma = 1 /$ incubation period |
| $\frac{dR}{dt} = +\gamma I$ | $\gamma = 1 /$ infectious period<br>$\tau = 1 /$ serial interval |

**Code and reproducibility.** The R scripts used to run the SEIR simulations and generate Fig 5 and the accompanying figures are publicly available at https://github.com/MarkYXY05/covid-lit-llm-code/tree/main/figures/seir. The parameter data file used for analysis is publicly available at https://github.com/MarkYXY05/covid-lit-llm-data. Final data package will be archived in Zenodo under the reserved DOI https://doi.org/10.5281/zenodo.22267835 before journal publication.

*Section 2. Comparison of AI- and human-extracted values*

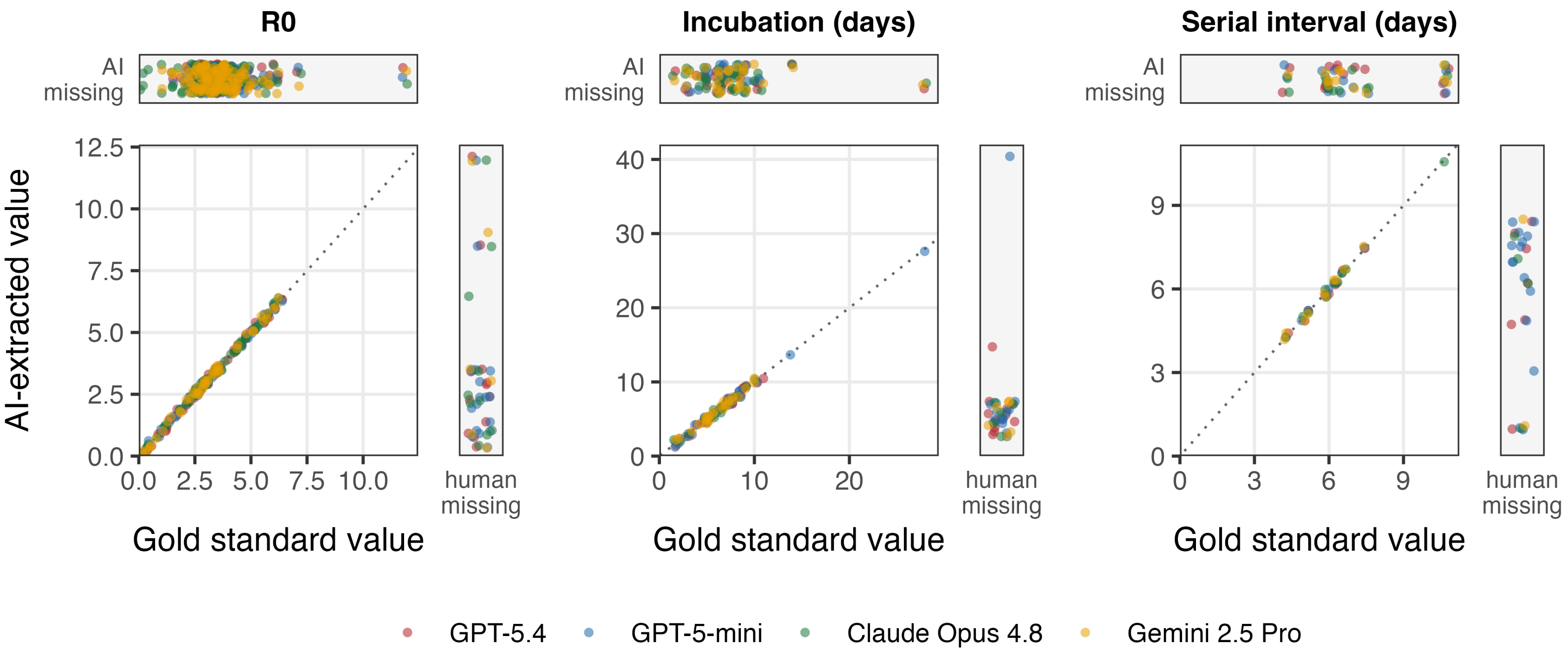


**Fig B.** ***Agreement between AI- and human-extracted values for basic reproduction number (R0), incubation period, and serial interval.*** *Each point is one extracted estimate, plotting the value extracted by an AI model (y-axis) against the corresponding human gold-standard value (x-axis), coloured by model: GPT-5.4 (red), GPT-5-mini (blue), Claude Opus 4.8 (green), and Gemini 2.5 Pro (yellow). The dotted line denotes perfect agreement (y = x); points on or near it were extracted identically to the gold standard. The top band ("AI missing") shows estimates humans extracted but the AI model did not (positioned by their gold-standard value only, with no y-axis value), and the right band ("human missing") shows estimates an AI model extracted but the humans did not (positioned by their AI value only, with no x-axis value). One high R0 value (59.3, correctly extracted by AI models and humans) was excluded for clarity.*

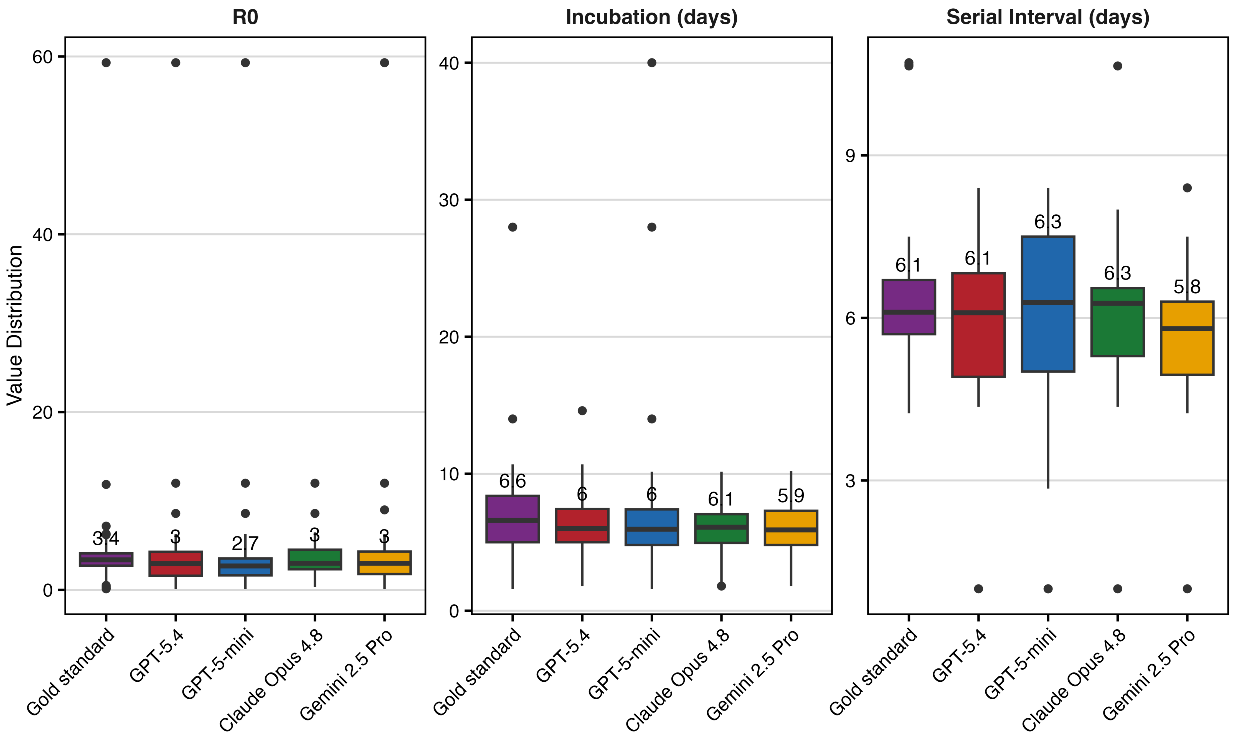


**Fig C. Boxplots showing the distribution of R0, incubation period, and serial interval for COVID-19 for extractions by human (purple), GPT-5.4 (red), GPT-5-mini (blue), Claude Opus 4.8 (green), and Gemini 2.5 Pro (yellow).** Of the 50 sampled studies, two meta-analyses were excluded to avoid double-counting. Of the remaining 48, 46 yielded at least one estimate of the three key parameters, from the human reviewers or from at least one model. Median values are labelled, and 25th and 75th quartiles, along with outliers, are shown.

### S7 Appendix. Full performance tables

#### *Section 1. Confusion counts*

**Table A. Full confusion-count master table: screening and value-level extraction (All, large-granularity-excluded, and extraction-only) for all four models.** Counts only; em-dash = not applicable to screening. The metrics derived from these counts are reported with confidence intervals in Table B for screening and in Table E for extraction.

| Stage | Model | Corpus | Scope | #AI | #Gold | TP | TN | FP | FN |
|---|---|---|---|---|---|---|---|---|---|
| Screening | GPT-5-mini | PubMed | — | — | — | 107 | 562 | 33 | 7 |
| Screening | GPT-5-mini | medRxiv | — | — | — | 369 | 893 | 75 | 20 |
| Screening | GPT-5.4 | PubMed | — | — | — | 94 | 589 | 6 | 20 |
| Screening | GPT-5.4 | medRxiv | — | — | — | 347 | 940 | 28 | 42 |
| Screening | Claude Opus 4.8 | PubMed | — | — | — | 91 | 582 | 13 | 23 |
| Screening | Claude Opus 4.8 | medRxiv | — | — | — | 281 | 955 | 13 | 108 |
| Screening | Gemini 2.5 Pro | PubMed | — | — | — | 100 | 583 | 12 | 14 |
| Screening | Gemini 2.5 Pro | medRxiv | — | — | — | 328 | 919 | 49 | 61 |
| Extraction | GPT-5-mini | PubMed | All | 58 | 80 | 53 | 32 | 5 | 27 |
| Extraction | GPT-5-mini | medRxiv | All | 115 | 240 | 109 | 28 | 6 | 131 |
| Extraction | GPT-5.4 | PubMed | All | 65 | 80 | 62 | 34 | 3 | 18 |
| Extraction | GPT-5.4 | medRxiv | All | 108 | 240 | 107 | 32 | 1 | 133 |
| Extraction | Claude Opus 4.8 | PubMed | All | 58 | 80 | 58 | 35 | 0 | 22 |
| Extraction | Claude Opus 4.8 | medRxiv | All | 94 | 240 | 91 | 31 | 3 | 149 |
| Extraction | Gemini 2.5 Pro | PubMed | All | 52 | 80 | 50 | 33 | 2 | 30 |
| Extraction | Gemini 2.5 Pro | medRxiv | All | 103 | 240 | 101 | 31 | 2 | 139 |
| Extraction | GPT-5-mini | PubMed | Excl-large | 53 | 52 | 48 | 32 | 5 | 4 |
| Extraction | GPT-5-mini | medRxiv | Excl-large | 79 | 84 | 73 | 28 | 6 | 11 |
| Extraction | GPT-5.4 | PubMed | Excl-large | 46 | 52 | 43 | 34 | 3 | 9 |
| Extraction | GPT-5.4 | medRxiv | Excl-large | 73 | 84 | 72 | 32 | 1 | 12 |
| Extraction | Claude Opus 4.8 | PubMed | Excl-large | 34 | 52 | 34 | 35 | 0 | 18 |
| Extraction | Claude Opus 4.8 | medRxiv | Excl-large | 64 | 84 | 61 | 31 | 3 | 23 |

| | | | | | | | | | |
|---|---|---|---|---|---|---|---|---|---|
| Extraction | Gemini 2.5 Pro | PubMed | Excl-large | 47 | 52 | 45 | 33 | 2 | 7 |
| Extraction | Gemini 2.5 Pro | medRxiv | Excl-large | 67 | 84 | 65 | 31 | 2 | 19 |
| Extraction | GPT-5-mini | PubMed | Extraction-only (excl-large) | 53 | 50 | 48 | 25 | 5 | 2 |
| Extraction | GPT-5-mini | medRxiv | Extraction-only (excl-large) | 79 | 84 | 73 | 28 | 6 | 11 |
| Extraction | GPT-5.4 | PubMed | Extraction-only (excl-large) | 46 | 46 | 43 | 23 | 3 | 3 |
| Extraction | GPT-5.4 | medRxiv | Extraction-only (excl-large) | 73 | 79 | 72 | 30 | 1 | 7 |
| Extraction | Claude Opus 4.8 | PubMed | Extraction-only (excl-large) | 34 | 37 | 34 | 20 | 0 | 3 |
| Extraction | Claude Opus 4.8 | medRxiv | Extraction-only (excl-large) | 64 | 66 | 61 | 20 | 3 | 5 |
| Extraction | Gemini 2.5 Pro | PubMed | Extraction-only (excl-large) | 47 | 50 | 45 | 26 | 2 | 5 |
| Extraction | Gemini 2.5 Pro | medRxiv | Extraction-only (excl-large) | 67 | 77 | 65 | 25 | 2 | 12 |

#### *Section 2. Screening performance*

**Table B. Screening performance against the human gold standard, by model and corpus (point estimate with 95% CI; operating point: uncertain = include).** Raw counts in Table A.

| Model | Corpus | Sensitivity | Specificity | Precision | F1 | AUC |
|---|---|---|---|---|---|---|
| GPT-5-mini | PubMed | 0.939 (0.88-0.97) | 0.945 (0.92-0.96) | 0.764 (0.69-0.83) | 0.843 | 0.973 (0.95-0.99) |
| GPT-5-mini | medRxiv | 0.949 (0.92-0.97) | 0.923 (0.90-0.94) | 0.831 (0.79-0.86) | 0.886 | 0.975 (0.97-0.98) |
| GPT-5.4 | PubMed | 0.825 (0.74-0.88) | 0.990 (0.98-0.99) | 0.940 (0.88-0.97) | 0.879 | 0.955 (0.93-0.98) |
| GPT-5.4 | medRxiv | 0.892 (0.86-0.92) | 0.971 (0.96-0.98) | 0.925 (0.89-0.95) | 0.908 | 0.971 (0.96-0.98) |
| Claude Opus 4.8 | PubMed | 0.798 (0.71-0.86) | 0.978 (0.96-0.99) | 0.875 (0.80-0.93) | 0.835 | 0.988 (0.98-0.99) |
| Claude Opus 4.8 | medRxiv | 0.722 (0.68-0.77) | 0.987 (0.98-0.99) | 0.956 (0.93-0.97) | 0.823 | 0.967 (0.96-0.97) |
| Gemini 2.5 Pro | PubMed | 0.877 (0.80-0.93) | 0.980 (0.96-0.99) | 0.893 (0.82-0.94) | 0.885 | 0.942 (0.91-0.97) |
| Gemini 2.5 Pro | medRxiv | 0.843 (0.80-0.88) | 0.949 (0.93-0.96) | 0.870 (0.83-0.90) | 0.856 | 0.914 (0.90-0.93) |

#### *Section 3. Ensemble performance*

**Table C. Screening k-of-N ensemble versus the best single model, full corpus (PubMed n = 709, 114 positives; medRxiv n = 1,357 of the 1,358 screened, 389 positives), against the human screening gold standard.** The ensemble vote-count ROC AUC is threshold-free (one value per corpus, shown on the union row); per-model figures appear in Table B. One medRxiv record was omitted when the ensemble was assembled; it is a gold-standard negative that all four models rejected, so no reported metric is affected.

| Corpus | Operating point | Sens. | Spec. | Prec. | F1 | F2 | AUC |
|---|---|---|---|---|---|---|---|
| PubMed | Best single (GPT-5-mini) | 0.939 | 0.945 | 0.764 | 0.843 | 0.898 | 0.973 |
| PubMed | Union (k≥1) | 0.965 | 0.924 | 0.710 | 0.818 | 0.900 | 0.993 |
| PubMed | k≥2 | 0.912 | 0.980 | 0.897 | 0.904 | 0.909 | — |
| PubMed | k≥3 | 0.851 | 0.992 | 0.951 | 0.898 | 0.869 | — |
| PubMed | Unanimous (k≥4) | 0.711 | 0.997 | 0.976 | 0.822 | 0.751 | — |
| medRxiv | Best single (GPT-5-mini) | 0.949 | 0.923 | 0.831 | 0.886 | 0.922 | 0.975 |
| medRxiv | Union (k≥1) | 0.982 | 0.904 | 0.804 | 0.884 | 0.940 | 0.987 |
| medRxiv | k≥2 | 0.910 | 0.948 | 0.876 | 0.893 | 0.903 | — |
| medRxiv | k≥3 | 0.807 | 0.979 | 0.940 | 0.869 | 0.831 | — |
| medRxiv | Unanimous (k≥4) | 0.707 | 0.998 | 0.993 | 0.826 | 0.750 | — |

**Table D. Extraction value-level support-voting ensemble versus the best single model, scored on the summary field (value matched together with its reported uncertainty), large-granularity entries excluded (the manuscript's extraction-only basis; PubMed 52, medRxiv 84 gold values).** Per-k audit detail is in the ensemble validation workbooks in the released data deposit.

| Corpus | Operating point | Recall | Precision | F1 | F2 |
|---|---|---|---|---|---|
| PubMed | Best single (GPT-5-mini) | 0.885 | 0.836 | 0.860 | 0.875 |
| PubMed | Union (k≥1) | 0.981 | 0.729 | 0.836 | 0.917 |
| PubMed | k≥2 | 0.885 | 0.868 | 0.876 | 0.881 |
| PubMed | k≥3 | 0.808 | 0.894 | 0.848 | 0.824 |
| PubMed | Unanimous (k≥4) | 0.423 | 0.917 | 0.579 | 0.474 |
| medRxiv | Best single (GPT-5.4) | 0.750 | 0.875 | 0.808 | 0.772 |
| medRxiv | Union (k≥1) | 0.845 | 0.710 | 0.772 | 0.814 |
| medRxiv | k≥2 | 0.762 | 0.800 | 0.780 | 0.769 |
| medRxiv | k≥3 | 0.702 | 0.908 | 0.792 | 0.736 |
| medRxiv | Unanimous (k≥4) | 0.429 | 0.947 | 0.590 | 0.481 |

#### *Section 4. Value-level extraction accuracy by scope*

Three scopes are reported for value-level extraction. Extraction-only and end-to-end both set aside articles that tabulate dozens of stratified values. Extraction-only scores each model on the articles it carried through screening, so screening exclusions are not charged against extraction. End-to-end scores every remaining gold value, so an article missed at screening contributes false negatives. The all scope retains the large-granularity articles. In the extraction-only results, precision ranged from 0.91 to 1.00, with Claude Opus 4.8 producing no false positives on PubMed, while GPT-5-mini achieved the highest recall there. Article-level exact-match accuracy was 0.67 to 0.93 and parameter-level accuracy was 0.85 to 1.00 across R0, incubation period, and serial interval.

**Table E. Value-level extraction quality by scope, model and corpus (point estimate with 95% CI).** Extraction-only scores only the articles a model carried through screening and excludes large-granularity entries; end-to-end scores every gold value on the same large-granularity-excluded basis, so articles missed at screening enter as false negatives; all retains the large-granularity entries as well. Precision is identical under the first two scopes because a screening exclusion adds false negatives without adding false positives, and its interval is given on the extraction-only row only. Intervals were computed for the two large-granularity-excluded scopes; all rows give point estimates derived from the counts in Table A. The screening cascade costs Claude Opus 4.8 the most on PubMed, where recall falls from 0.919 to 0.654, and costs GPT-5-mini nothing on medRxiv.

| Model | Corpus | Scope | #Gold | Recall (95% CI) | Precision (95% CI) | F1 (95% CI) | F2 |
|---|---|---|---|---|---|---|---|
| GPT-5-mini | PubMed | Extraction-only | 50 | 0.960 (0.87-1.00) | 0.906 (0.80-0.96) | 0.932 (0.85-0.99) | 0.949 |
| GPT-5-mini | PubMed | End-to-end | 52 | 0.923 (0.79-1.00) | 0.906 | 0.914 | 0.920 |
| GPT-5-mini | PubMed | All | 80 | 0.662 | 0.914 | 0.768 | 0.701 |
| GPT-5-mini | medRxiv | Extraction-only | 84 | 0.869 (0.74-0.99) | 0.924 (0.84-0.97) | 0.896 (0.81-0.97) | 0.880 |
| GPT-5-mini | medRxiv | End-to-end | 84 | 0.869 (0.74-0.99) | 0.924 | 0.896 | 0.880 |
| GPT-5-mini | medRxiv | All | 240 | 0.454 | 0.948 | 0.614 | 0.507 |
| GPT-5.4 | PubMed | Extraction-only | 46 | 0.935 (0.82-1.00) | 0.935 (0.83-0.98) | 0.935 (0.84-1.00) | 0.935 |
| GPT-5.4 | PubMed | End-to-end | 52 | 0.827 (0.65-0.96) | 0.935 | 0.878 | 0.846 |
| GPT-5.4 | PubMed | All | 80 | 0.775 | 0.954 | 0.855 | 0.805 |
| GPT-5.4 | medRxiv | Extraction-only | 79 | 0.911 (0.80-1.00) | 0.986 (0.93-1.00) | 0.947 (0.89-1.00) | 0.925 |
| GPT-5.4 | medRxiv | End-to-end | 84 | 0.857 (0.70-0.99) | 0.986 | 0.917 | 0.880 |
| GPT-5.4 | medRxiv | All | 240 | 0.446 | 0.991 | 0.615 | 0.501 |
| Claude Opus 4.8 | PubMed | Extraction-only | 37 | 0.919 (0.79-1.00) | 1.000 (0.90-1.00) | 0.958 (0.88-1.00) | 0.934 |
| Claude Opus 4.8 | PubMed | End-to-end | 52 | 0.654 (0.42-0.86) | 1.000 | 0.791 | 0.702 |
| Claude Opus 4.8 | PubMed | All | 80 | 0.725 | 1.000 | 0.841 | 0.767 |
| Claude Opus 4.8 | medRxiv | Extraction-only | 66 | 0.924 (0.79-1.00) | 0.953 (0.87-0.98) | 0.938 (0.86-1.00) | 0.930 |
| Claude Opus 4.8 | medRxiv | End-to-end | 84 | 0.726 (0.53-0.91) | 0.953 | 0.824 | 0.762 |
| Claude Opus 4.8 | medRxiv | All | 240 | 0.379 | 0.968 | 0.545 | 0.432 |
| Gemini 2.5 Pro | PubMed | Extraction-only | 50 | 0.900 (0.78-1.00) | 0.957 (0.86-0.99) | 0.928 (0.86-0.99) | 0.911 |
| Gemini 2.5 Pro | PubMed | End-to-end | 52 | 0.865 (0.74-1.00) | 0.957 | 0.909 | 0.882 |
| Gemini 2.5 Pro | PubMed | All | 80 | 0.625 | 0.962 | 0.758 | 0.672 |
| Gemini 2.5 Pro | medRxiv | Extraction-only | 77 | 0.844 (0.73-0.96) | 0.970 (0.90-0.99) | 0.903 (0.83-0.97) | 0.867 |
| Gemini 2.5 Pro | medRxiv | End-to-end | 84 | 0.774 (0.62-0.92) | 0.970 | 0.861 | 0.806 |
| Gemini 2.5 Pro | medRxiv | All | 240 | 0.421 | 0.981 | 0.589 | 0.475 |

### S8 Appendix. Operational guidance and what we tested

Table A summarizes safeguards used in the locked pipeline and changes considered after the error analysis. Status refers to implementation during the reported main runs unless a post hoc analysis is specified. Prompt-only safeguards were instructions that are stated in model prompts but not enforced by the schema or downstream code. Partial implementation means that relevant information was produced without a complete operational check.

**Table A. Safeguards considered in the study and their implementation status.**

| Workflow safeguard | Status in locked pipeline | Evidence from this study | Implication for future use |
|---|---|---|---|
| Recall-oriented screening | Implemented in all reported runs. | Uncertain decisions advanced to extraction; excluded articles contributed downstream false negatives. | A recall-oriented screening threshold remains appropriate. |
| Low-certainty human routing | Not implemented prospectively; assessed post hoc. | For Claude Opus 4.8, reviewing decisions at certainty ≤7 recovered 58 of 131 missed articles (44%) at an 8% review burden. Certainty did not separate errors for the other models. | Certainty would require model-specific calibration before it could guide routing. |
| Targeted human verification of extracted values | Not evaluated as a prospective routing strategy. | Extraction precision was 0.91–1.00, and no fabricated values were observed. | Targeted review of borderline and provenance-sensitive outputs requires prospective evaluation. |
| Structured provenance fields | Recorded in narrative summaries but not enforced structurally. | Models often labelled values as assumed or cited but still placed them in scored estimate fields. | Structured provenance fields could support parsing and validation. |
| Per-value pathogen attribution | Present as a prompt instruction; not enforced in the schema. | An influenza R0 was placed in the COVID-19 R0 field although the summary identified the pathogen. | Pathogen labels should be recorded and checked for each value. |
| Separate fields for related parameters | Not implemented in the locked pipeline. | R0 and Rt, as well as serial interval and generation time, were sometimes conflated and required manual separation. | Separate schema fields could reduce manual separation of related quantities. |
| Few-shot boundary examples | Not implemented or evaluated. | The locked prompt contained formatting examples but no boundary examples. | Boundary examples could be tested in future prompt designs. |
| One-row-per-estimate output | Not implemented in the locked pipeline; evaluated post hoc in S4 Appendix. | The alternative row-based configuration recovered more dense-target values, but precision decreased under some conditions (S4 Appendix, Tables A and C). | For dense articles, row-wise output warrants further evaluation with precision monitoring. |
| Downstream provenance filtering | Provenance labels were generated; automated filtering was not implemented. | The summaries contained provenance information, but no filter was built or evaluated. | Any provenance-based exclusion filter would require separate validation. |

### S9 Appendix. Efficiency and cost

**Table A. Computational cost of the full pipeline by model, with two human annotators shown for comparison. Automated totals cover screening of the 2,065 records with complete cost logs and full-text extraction of records passing screening. Token counts are exact. API costs are approximate estimates anchored to billed spend, which also included development and re-runs. Model time is the median per-article sum of API-call latencies; human time is the mean annotation time across 50 timed gold-standard articles. Concurrent model calls reduced total wall-clock time to a few hours. Values plotted in Fig A are from this table.**

| Model | Total tokens | Tokens / article | Time per article | Approx. API cost |
|---|---|---|---|---|
| GPT-5-mini (deployed) | 36M | ~17,500 | 9.5 s | ~$25 |
| GPT-5.4 | 29M | ~14,100 | 3.4 s | ~$60–90 |
| Claude Opus 4.8 | 40M | ~19,200 | 4.4 s | ~$175–240 |
| Gemini 2.5 Pro | 37M | ~18,000 | 10.6 s | ~$70–120 |
| Human annotator 1 | — | — | 8.9 min (PubMed); 14.4 min (medRxiv) | — |
| Human annotator 2 | — | — | 19.6 min (PubMed); 24.5 min (medRxiv) | — |

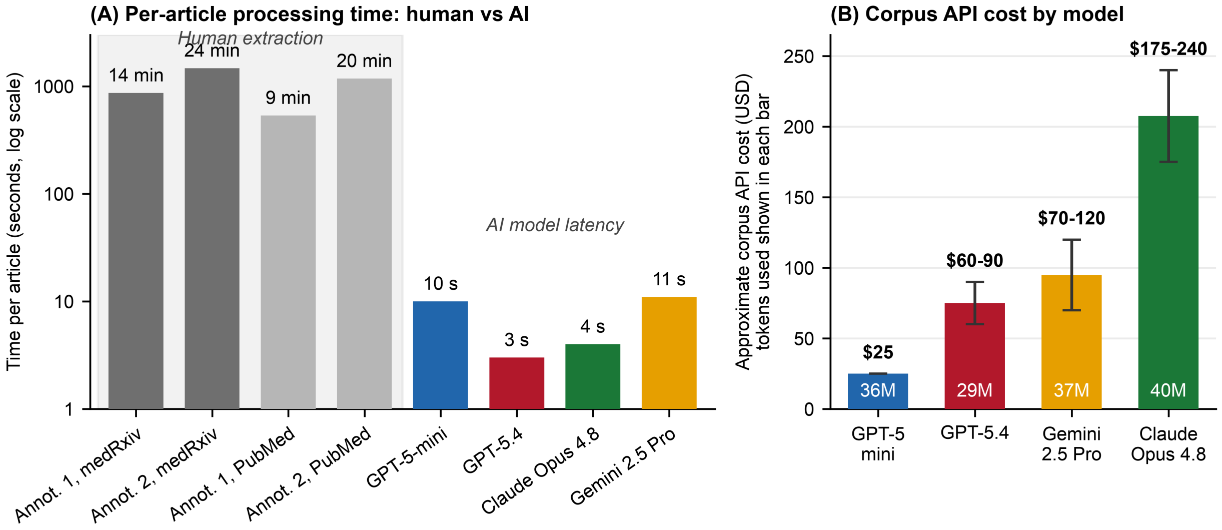


**Fig A. Computational cost of automated extraction and manual effort. (A) Per-article processing time for human extraction (minutes; two annotators × two corpora) and model latency (median seconds; four models), shown on a logarithmic scale. (B) Approximate corpus-scale API cost and token use by model. Token counts and cost estimates are given in Table A.**

PRISMA

### PRISMA 2020 Checklist

**S10 Checklist. PRISMA 2020 checklist.**

| Section and Topic | Item # | Checklist item | Location where item is reported |
|---|---|---|---|
| **TITLE** | | | |
| Title | 1 | Identify the report as a systematic review. | Not applicable. This report evaluates an automated screening and extraction pipeline; it is not itself a systematic review report. The review described here supplies the human gold standard against which the pipeline was scored. |
| **ABSTRACT** | | | |
| Abstract | 2 | See the PRISMA 2020 for Abstracts checklist. | Abstract. Registration and certainty-of-evidence items do not apply. |
| **INTRODUCTION** | | | |
| Rationale | 3 | Describe the rationale for the review in the context of existing knowledge. | Introduction. |
| Objectives | 4 | Provide an explicit statement of the objective(s) or question(s) the review addresses. | Introduction, final paragraph. |
| **METHODS** | | | |
| Eligibility criteria | 5 | Specify the inclusion and exclusion criteria for the review and how studies were grouped for the syntheses. | Methods, Human gold standard. Full inclusion and exclusion criteria in S1 Appendix, Section 2. |
| Information sources | 6 | Specify all databases, registers, websites, organisations, reference lists and other sources searched or consulted to identify studies. Specify the date when each source was last searched or consulted. | Methods, Database search. PubMed and medRxiv, 31 December 2019 to 30 April 2020. Search dates and retrieval route for each source in S1 Appendix, Section 3. |
| Search strategy | 7 | Present the full search strategies for all databases, registers and websites, including any filters and limits used. | S1 Appendix, Section 1 gives the full PubMed and medRxiv search strings with their date limits; Section 3 gives the date each search was run. |
| Selection process | 8 | Specify the methods used to decide whether a study met the inclusion criteria of the review, including how many reviewers screened each record and each report retrieved, whether they worked independently, and if applicable, details of automation tools used in the process. | Methods, Human gold standard. One reviewer screened, with uncertain records re-adjudicated by a blinded expert reviewer. Details noted in S1 Appendix, Section 4, Fig B. |
| Data collection process | 9 | Specify the methods used to collect data from reports, including how many reviewers collected data from each report, whether they worked independently, any processes for obtaining or confirming data from study investigators, and if applicable, details of automation tools used in the process. | Methods, Human gold standard. Two reviewers extracted independently and blinded to model output; discrepancies reconciled by discussion and expert adjudication. Written protocol in S1 Appendix, Section 3. |
| Data items | 10a | List and define all outcomes for which data were sought. Specify whether all results that were compatible with each outcome domain in each study were sought (e.g. for all measures, time points, analyses), and if not, the methods used to decide which results to collect. | Methods, Human gold standard and LLM-facilitated pipeline. The reproduction number, serial interval and incubation period, each as a point estimate with its reported uncertainty. |
| | 10b | List and define all other variables for which data were sought (e.g. participant and intervention characteristics, funding sources). Describe any assumptions made about any missing or unclear information. | Methods, LLM-facilitated pipeline. Study design, geography, time window, sample, subgroup, provenance of each value and verbatim supporting quotations. Full schema in S2 Appendix. |
| Study risk of | 11 | Specify the methods used to assess risk of bias in the included studies, including details of the tool(s) | Not applicable. Risk of bias in the included studies |

### PRISMA 2020 Checklist

| Section and Topic | Item # | Checklist item | Location where item is reported |
|---|---|---|---|
| bias assessment | | used, how many reviewers assessed each study and whether they worked independently, and if applicable, details of automation tools used in the process. | was not assessed. The quantities pooled here are epidemiological parameter values rather than intervention effect estimates, and the object of evaluation is the extraction pipeline rather than the evidence base. |
| Effect measures | 12 | Specify for each outcome the effect measure(s) (e.g. risk ratio, mean difference) used in the synthesis or presentation of results. | Methods, Pooled estimates and heterogeneity. No effect measure applies: the quantities synthesised are parameter point estimates with their reported uncertainty intervals. |
| Synthesis methods | 13a | Describe the processes used to decide which studies were eligible for each synthesis (e.g. tabulating the study intervention characteristics and comparing against the planned groups for each synthesis (item #5)). | Methods, Pooled estimates and heterogeneity. Only estimates carrying a 95% CI, CrI or eCI, or an SD convertible to one, entered a synthesis. |
| | 13b | Describe any methods required to prepare the data for presentation or synthesis, such as handling of missing summary statistics, or data conversions. | Methods, Pooled estimates and heterogeneity. SD converted as mean plus or minus 1.96 x SD; two meta-analyses excluded to avoid double-counting. |
| | 13c | Describe any methods used to tabulate or visually display results of individual studies and syntheses. | Results, Pooled parameter estimates and transmission model. Raw value distributions and forest plots in S6 Appendix. |
| | 13d | Describe any methods used to synthesize results and provide a rationale for the choice(s). If meta-analysis was performed, describe the model(s), method(s) to identify the presence and extent of statistical heterogeneity, and software package(s) used. | Methods, Pooled estimates and heterogeneity. Inverse-variance weighted random-effects model, with each study's estimates weighted 1/n. |
| | 13e | Describe any methods used to explore possible causes of heterogeneity among study results (e.g. subgroup analysis, meta-regression). | Methods, Pooled estimates and heterogeneity. Tau squared, I squared with 95% CI, and Cochran's Q. |
| | 13f | Describe any sensitivity analyses conducted to assess robustness of the synthesized results. | Methods, Sensitivity analysis. Note that this varies the task specification, namely output format, instruction wording, input mode and aggregation, rather than the synthesis model. No sensitivity analysis of the meta-analytic model was conducted. |
| Reporting bias assessment | 14 | Describe any methods used to assess risk of bias due to missing results in a synthesis (arising from reporting biases). | Not applicable. Reporting bias was not assessed. The corpus comprised records retrieved within a predefined four-month window rather than a sample, so funnel-plot and regression-based tests are not applicable for this pipeline evaluation design. |
| Certainty assessment | 15 | Describe any methods used to assess certainty (or confidence) in the body of evidence for an outcome. | Not applicable. Certainty of evidence was not graded. The study evaluates extraction accuracy against a human reference standard and draws no clinical or policy conclusion from the pooled parameters. |
| **RESULTS** | | | |
| Study selection | 16a | Describe the results of the search and selection process, from the number of records identified in the search to the number of studies included in the review, ideally using a flow diagram. | S1 Appendix, Section 4: Fig A for the human gold-standard review, Fig B for the automated arm. Counts also in Results, Literature search and automated screening performance. |

PRISMA

### PRISMA 2020 Checklist

| Section and Topic | Item # | Checklist item | Location where item is reported |
|---|---|---|---|
| | 16b | Cite studies that might appear to meet the inclusion criteria, but which were excluded, and explain why they were excluded. | Results, Literature search and automated screening performance. Exclusions by criterion in S3 Appendix. |
| Study characteristics | 17 | Cite each included study and present its characteristics. | Partially. The 50 articles of the extraction gold standard are characterised in S6 Appendix and in the released data. The 503 included records are not tabulated individually. |
| Risk of bias in studies | 18 | Present assessments of risk of bias for each included study. | Not applicable. See item 11. |
| Results of individual studies | 19 | For all outcomes, present, for each study: (a) summary statistics for each group (where appropriate) and (b) an effect estimate and its precision (e.g. confidence/credible interval), ideally using structured tables or plots. | Per-article, per-parameter results for the 50 gold-standard articles are in the released data; S6 Appendix, Fig B plots every extracted value against its gold-standard counterpart. |
| Results of syntheses | 20a | For each synthesis, briefly summarise the characteristics and risk of bias among contributing studies. | Results, Pooled parameter estimates and transmission model; Table 1. |
| | 20b | Present results of all statistical syntheses conducted. If meta-analysis was done, present for each the summary estimate and its precision (e.g. confidence/credible interval) and measures of statistical heterogeneity. If comparing groups, describe the direction of the effect. | Results, Pooled parameter estimates and transmission model; Table 1. |
| | 20c | Present results of all investigations of possible causes of heterogeneity among study results. | Results, Pooled parameter estimates and transmission model; Table 1 reports tau squared, I squared and Cochran's Q. |
| | 20d | Present results of all sensitivity analyses conducted to assess the robustness of the synthesized results. | Not applicable. No sensitivity analysis of the synthesis was conducted. The sensitivity analysis reported in Results concerns the task specification; see item 13f. |
| Reporting biases | 21 | Present assessments of risk of bias due to missing results (arising from reporting biases) for each synthesis assessed. | Not applicable. See item 14. |
| Certainty of evidence | 22 | Present assessments of certainty (or confidence) in the body of evidence for each outcome assessed. | Not applicable. See item 15. |
| **DISCUSSION** | | | |
| Discussion | 23a | Provide a general interpretation of the results in the context of other evidence. | Discussion, Principal findings; Relation to Prior Work and What This Study Adds. |
| | 23b | Discuss any limitations of the evidence included in the review. | Discussion, Strengths, limitations and recommendations for future work. |
| | 23c | Discuss any limitations of the review processes used. | Discussion, Strengths, limitations and recommendations for future work. |
| | 23d | Discuss implications of the results for practice, policy, and future research. | Discussion, Practical guidance for specifying a workflow. |
| **OTHER INFORMATION** | | | |
| Registration and protocol | 24a | Provide registration information for the review, including register name and registration number, or state that the review was not registered. | Methods, Database search. The review was not registered. |

PRISMA

### PRISMA 2020 Checklist

| Section and Topic | Item # | Checklist item | Location where item is reported |
|---|---|---|---|
| | 24b | Indicate where the review protocol can be accessed, or state that a protocol was not prepared. | Methods, Database search. No protocol was posted to a registry or published separately. The search strategy, the screening criteria and the gold-standard extraction protocol are reproduced in S1 Appendix. |
| | 24c | Describe and explain any amendments to information provided at registration or in the protocol. | Not applicable. There was no registration or published protocol to amend. The screening criteria and the extraction pipeline were locked before the evaluation runs and were not changed thereafter. |
| Support | 25 | Describe sources of financial or non-financial support for the review, and the role of the funders or sponsors in the review. | Financial Disclosure, provided in the submission system. |
| Competing interests | 26 | Declare any competing interests of review authors. | Competing Interests. |
| Availability of data, code and other materials | 27 | Report which of the following are publicly available and where they can be found: template data collection forms; data extracted from included studies; data used for all analyses; analytic code; any other materials used in the review. | Data and Code Availability. |

*From:* Page MJ, McKenzie JE, Bossuyt PM, Boutron I, Hoffmann TC, Mulrow CD, et al. The PRISMA 2020 statement: an updated guideline for reporting systematic reviews. BMJ 2021;372:n71. doi: 10.1136/bmj.n71. This work is licensed under CC BY 4.0. To view a copy of this license, visit https://creativecommons.org/licenses/by/4.0/
